# A circuit-based approach to repair hypersexuality in Parkinson’s disease

**DOI:** 10.1101/2022.01.25.22269798

**Authors:** David Mata-Marín, José Ángel Pineda-Pardo, Mario Michiels, Cristina Pagge, Claudia Ammann, Raúl Martínez-Fernández, José Antonio Molina, Lydia Vela-Desojo, Fernando Alonso-Frech, Ignacio Obeso

## Abstract

One common neuropsychiatric complication in patients with Parkinson’s disease treated with dopamine replacement therapy is impulse control disorders. A proportion of patients under dopamine agonist exhibit excessive desire towards appetitive cues accompanied by lost control over behaviour in forms of pathological gambling, hypersexuality or binge eating. The balance between enhanced desire towards sexual cues and cognitive control changes has been hypothesized to be biased toward the former in individuals with hypersexuality and impulse control disorders. Yet, some studies report no behavioural differences in cognitive control differences between impulsive patients and general Parkinson’s disease, suggesting possible functional changes along the mesocorticolimbic cortico-subcortical circuitry. Here, we provide evidence for this hypothesis by comparing the neurobiological substrate of sexual disturbance over response inhibition associated to medication states in Parkinson’s disease patients with a specific subtype of impulse control disorders (i.e., hypersexuality).

We assessed the impact of sexual cues on response inhibition using a novel erotic stop signal task inside an fMRI. A total of 50 participants were included divided in 16 hypersexual and 17 non-hypersexual Parkinson’s disease patients and 17 healthy controls. Task-related activations, functional and anatomical connectivity models were performed. Additionally, a separate sample of 20 hypersexual Parkinson’s disease patients received excitatory neuromodulation (sham-controlled) over the pre-supplementary motor area (based on fMRI group-based results) aiming to improve response inhibition when exposed to sexual cues.

Compared with their non-hypersexual peers, medication perturbed response inhibition upon presentation of sexual cues in patients with hypersexuality, recruiting a network involving caudate, pre-supplementary motor area, ventral tegmental area, and anterior cingulate cortex. Dynamic causal modelling revealed distinct best models to account for cortico-subcortical interactions with reduced task-related inputs in pre-supplementary motor area and descending connectivity to caudate in hypersexual compared to non- hypersexual Parkinson’s disease patients (while medicated). This was sustained by enhanced fractional anisotropy and reduced mean diffusivity in the pre-supplementary motor area-caudate pathway. Importantly, stimulation over the pre-supplementary motor area improved response inhibition when exposed to sexual cues in hypersexual Parkinson’s disease.

We identified a specific fronto-striatal and mesolimbic circuitry underlying uncontrolled sexual behaviours in Parkinson’s disease induced by medication, with recovery options by applying neuromodulation.

## Introduction

Impulse control disorder (ICD) corresponds to one of the most acute and disturbing neuropsychiatric complication in Parkinson’s disease patients under dopaminergic replacement therapy. The most prevalent forms include compulsive shopping, pathological gambling, sexual behaviours or binge-eating amongst others, all marked by the excessive tendency to compulsively engage in rewarding uncontrolled actions. ICDs are seen in about 17% of Parkinson’s disease patients induced both by dopamine agonist and more rarely by levodopa (6.9%)^1–4^. ICD sets patients at increased risk of financial ruin, family problems, prosecution, or job-related problems. Yet, its treatment follows two clinical routes: reducing dopaminergic doses at the expense of worsening movement output or invasive surgery. Hence, no specific treatments are available in ICD, prompting the available therapeutic windows to be consumed earlier than expected ^5^.

It is commonly accepted that ICD is the result of a dopaminergic dysregulation of the mesolimbic pathway – including the orbital prefrontal cortex, anterior cingulate cortex (ACC), ventral striatum (nucleus accumbens), amygdala and ventral tegmental area (VTA) – that mediates reward. Mesolimbic dopaminergic dysregulation is thought to result from a hyperdopaminergic state caused by the action of dopaminergic therapy in the lesser denervated limbic striatum compared to more affected motor territories of the striatum^6^. Indeed, the dopaminergic dysregulation changes reported in ICD along the mesolimbic pathway are similar to those seen in addiction pathophysiology^7, 8^. As a result, enhanced desire towards appetitive items occurs in ICD, driving excessive approach behaviours that find little or no opposition.

A dysregulated limbic system in Parkinson’s disease and ICD sets excessive desire to primary and/or secondary rewards, possibly interfering with the neurocognitive signature needed to apply cognitive control operations. Indeed, Parkinson’s disease and ICD patients reveal larger tendency for novelty seeking^9^ and enhanced incentive salience^10,11^, compatible with views on altered reward processing. Regarding cognitive control, Parkinson’s disease and ICD modifies set-shifting abilities^12–14^ and impairs adaptation from negative outcomes^15–17^. Moreover, these impulsive patients show preference for smaller immediate rewards over larger and delayed ones^18,19^. Yet, we are uncertain whether a lack of cognitive control, enhanced desire or their imbalance is the basis of ICD associated to mesocorticolimbic regions.

The “controlling brain network” in humans (namely inferior frontal gyrus, pre-supplementary motor area (pre-SMA), striatum and the subthalamic nucleus^20–24^ is underactive in Parkinson’s disease and ICD patients when making monetary choices, partly clarifying the clinical problem showing uncontrolled behaviour^25,26^. Yet, unaltered response inhibition in Parkinson’s disease and ICD patients^27,28^ contrasts with altered engagement of the controlling network^29,30^ and increased engagement of the salience brain network responsible for switching between different brain neural dynamics^31,32^. Most studies include mixed ICD subtypes and evaluate behaviour with stimuli unrelated to their impulsive problem. Thus, to reduce the potential bias of unrelated stimuli with patient’s impulsivity, the study of one predominant ICD using ad-hoc provocative stimuli will better pinpoint the specific circuits part of the clinical issue.

To this, we recruited Parkinson’s disease patients with hypersexuality (PD+HS) and designed a novel paradigm combining erotic cues within a stop-signal task to investigate the behavioural, neural and connectivity (anatomical and functional) profile in PD+HS (study 1). Hypersexuality is one of the most common subtypes of ICD with an estimated prevalence of 13.6% in Parkinson’s disease^2^. Hence, given response inhibition is vulnerable to high-arousal stimuli^33,34^, we hypothesized that sexual cues will bias response inhibition by altering fronto-striatal networks in hypersexual Parkinson’s disease patients (PD+HS). Next, we hypothesized that response inhibition in PD+HS patients can be improved by means of repetitive transcranial magnetic stimulation (rTMS, study 2) – a non-invasive neuromodulation technique effective in improving response inhibition in the general and neuropsychiatric population^35–37^. Our design represents an advantage over prior studies as it counts with one ICD sub-type and assessed on response inhibition performance using ad hoc stimuli to provoke their symptomatology. The circuit-based approach hereby utilized may serve to elucidate the neurobiological changes when response inhibition is influenced by sexual desire in a specific subtype of ICD and use possible treatment options to offer novel therapies to a yet untreatable complication.

## Materials and methods

### Participants

Patients in both studies were heterosexual male and right-handed. In study 1, we included 50 participants divided into 16 PD+HS patients, 17 non-hypersexual Parkinson’s disease patients (PD-HS) and 17 controls (**Table 1**). Study 2 included 20 PD+HS patients. Both studies are consistent with sample sizes used in previous neuroimaging and stimulation studies in Parkinson’s disease and ICD^35,38^. Patients met UK Brain Bank criteria for Parkinson’s disease diagnosis. Due to the special recruitment criteria, 4 hospitals participated in the recruitment. Recruitment was confirmed after an initial interview with the neuropsychologist (with both patient and families) to confirm heterosexuality and hypersexuality (≥5 on the sexual subscale of the Questionnaire for Impulsive-Compulsive Disorder in Parkinson’s disease). The PD-HS group was selected based on absent or limited presence of ICD-related behaviours, while on-medication (<4 on the sexual subscale of the Questionnaire for Impulsive-Compulsive disorder in Parkinson’s disease). Exclusion criteria were established based on presence of (i) cognitive dysfunction (≤24 on the Montreal Cognitive Assessment) (ii) depression (>18 in Beck Depression Inventory) or (iii) other severe comorbidities (e.g., history of substance abuse, homosexuality, hallucinations, or psychosis) detected during the interview. The study was approved by the Hospital Ethics Committee (17.03.0852E2-GHM).

**Table 1.** Demographic, clinical, and neuropsychological data for each group.

|  | Study 1 |  |  |  | Study 2 |
| --- | --- | --- | --- | --- | --- |
|  | PD+HS<br>(n=14) | PD-HS<br>(n=14) | Controls<br>(n=16) | Stats* | PD+HS<br>(n=18) |
| *Age* | 62.50±9.1 | 65.40±5.4 | 58.12±9.8 | .96 | 57.89±8.0 |
| *Education* | 12.50±4.3 | 12.93±3.9 | 14.35±3.4 | .69 | 12.71±2.9 |
| *Disease duration* | 8.38±3.2 | 6.27±3.9 | - | .14 | 6.89±4.5 |
| *UPDRS-III<sup>a</sup>*<br>(OFF) | 29.31±7.4 | 27.80±10.4 | - | .41 | 28.65±11.4 |
| *UPDRS-III (ON)* | 15.13±5.1 | 14.53±6.2 | - | .73 | 17.47±7.71 |
| *LEDD<sup>b</sup> (total)* | 740.63±273.3 | 549.29±293.2 | - | .12 | 738.61±631.2 |
| *LEDD<sup>c</sup> (DA)* | 206.67±84.3 | 171.82±76.4 | - | .44 | 362.67±74.3 |
| *LEDD<sup>d</sup> (L-dopa)* | 583.33±237.3 | 527.27±262.0 | - | .52 | 375.94±286.8 |
| *QUIP<sup>e</sup> total* | 16.24±11.9 | 5.27±8.5 | 7.88±10.6 | <.01 | 30.89±14.99 |
| Hypersexual | 6.41±3.9 | 1.20±1.7 | 1.47±1.8 | <.001 | 8.61±3.78 |
| Gambling | 0.65±1.2 | 0.20±0.5 | 0.29±0.9 | .21 | 1.00±2.17 |
| Shopping | 1.76±2.3 | 0.67±1.4 | 1.47±1.7 | .12 | 3.83±4.45 |
| Binge eating | 2.76±3.3 | 0.67±1.1 | 1.46±1.9 | .02 | 4.33±2.99 |
| Hobbyism | 1.35±2.2 | 1.00±1.9 | 1.65±2.3 | .63 | 4.56±4.02 |
| Pounding | 1.24±2.3 | 0.67±1.8 | 1.47±2.3 | .45 | 4.11±3.72 |
| Medication | 2.12±3.6 | 0.87±1.7 | 0.06±0.2 | .23 | 4.44±4.08 |
| *MoCa<sup>f</sup>* | 27.67±1.5 | 28.00±1.3 | 27.42±1.2 | .41 | 27.94±2.1 |
| *Stroop* | 79.6±37.3 | 62.6±14.5 | 51.18±16.5 | .10 | 57.89±8.1 |
| *FAB<sup>g</sup>* | 17.33±0.9 | 16.93±1.8 | 17.94±0.2 | .24 | 16.88±1.8 |
| *GDS<sup>h</sup>* | 7.92±4.4 | 8.77±6.1 | 3.59±2.4 | .59 | 8.18±7.1 |
| *BAI<sup>i</sup>* | 11.85±5.0 | 14.69±8.5 | 6.94±4.3 | .29 | 12.06±10.0 |
| *SAS<sup>j</sup>* | 10.08±4.7 | 12.15±8.8 | 4.29±4.6 | .46 | 11.94±6.4 |
| *BIS-III<sup>k</sup>* | 79.50±18.5 | 73.69±12.6 | 81.08±12.8 | .39 | 58.44±8.4 |
| *BSFI<sup>l</sup>* | 26.82±10.2 | 21.00±7.1 | 23.75±12.5 | .11 | 12.71±2.9 |

Stats: unpaired t-test between PD+HS and PD-HS; between groups; <sup>a</sup>UPDRS-III: Unified Parkinson's Disease Rating Scale;<sup>b</sup>LEDD: Levodopa Equivalent Daily Dose (total mg). <sup>c</sup>LEDD-DA: Levodopa Equivalent Daily Dose of Dopamine Agonist (total DA mg); <sup>d</sup>LEDD-L-dopa: Levodopa Equivalent Daily Dose (total Levodopa mg); <sup>e</sup>QUIP: Questionnaire for Impulsive-Compulsive;<sup>f</sup>MoCa: Montreal Cognitive Assessment; <sup>g</sup>FAB: Frontal Assessment Battery; <sup>h</sup>GDS: Geriatric Depression Scale; <sup>i</sup>BAI: Beck's Anxiety Inventory; <sup>j</sup>SAS: Starkstein Apathy Scale; Disorders in Parkinson's; <sup>k</sup>BIS-III: Barratt Impulsiveness Scale; <sup>l</sup>BSFI: Brief Sexual Function Inventory.

### Procedure

Patients were assessed while on and off-medication states (study 1, off-state defined as overnight dopaminergic drug withdrawal ∼12hs) inside the functional MRI (fMRI) scanner with at least one week time between each session (order counterbalanced). Scores of internal arousal state were collected to obtain explicit measurements of sexual desire – by asking participants to mark between 0 (nothing) to 10 (very much) on how intense their internal state for pre-scanning was:

1. How much sexual appetite did you experienced in the last hour?
2. Did you feel increased libido in the last hour? Similarly, after the fMRI session, in addition to the first 2 items above, the following items were included:
3. How much you felt nude images corrupted or slowed your responses?
4. How much you felt dressed images corrupted or slowed your responses?

Behavioural measures (classic stop signal task), neuropsychological, and neuropsychiatric tests were performed while on medication in both studies.

A single-blinded randomized controlled trial was designed to evaluate the acute effects of rTMS (study 2). Each patient underwent both real and sham stimulation sessions in separate two days maximum two-weeks apart (counterbalanced; on medication). Following stimulation, patients performed the erotic stop-signal task described below.

### The erotic stop-signal task

We designed an erotic stop-signal task using sexual cues (2 parallel versions; using a previous database^39^ and additional images). Erotic and non-erotic images (added to Prevost *et al.*^39^) were rated in a pre-experiment pilot phase, were healthy heterosexual male participants (*n*=22; *age (mean*±*sd*=48.4±2.5) rated valence and arousal states of the new images. We selected the most consistent high-valued images to insert in the task design. Images of undressed (erotic) and dressed women (non-erotic) were displayed followed by left or right-pointing arrows (**Figure 1A**), where a left (index finger) or right keypress (middle finger) with their dominant hand (limited time-hold up to 1s) was required. On Stop trials (33%), a stop signal cue was presented after a variable stop signal delay (staircase procedure using 50ms steps) following go signals, asking patients to stop their ongoing response. Null events ranged between 3-5s. Measures included response initiation, stop signal reaction time (SSRT; integration method by replacing go omissions with the maximum reaction time values per participant^40^, stop signal delay and proactive action restraint as response delay effect (difference between erotic and non-erotic go reaction times). Go trials above 3.5 standard deviation from the participant’s mean reaction time were discarded from the analysis. Twenty practice trials outside the scanner were performed before each scanning session. A total of 384 trials (192 per image condition) were performed per session, divided in five blocks of 77 trials per block. Stimulus presentation and response recording were controlled using E-prime 2.0.

**Figure 1.**
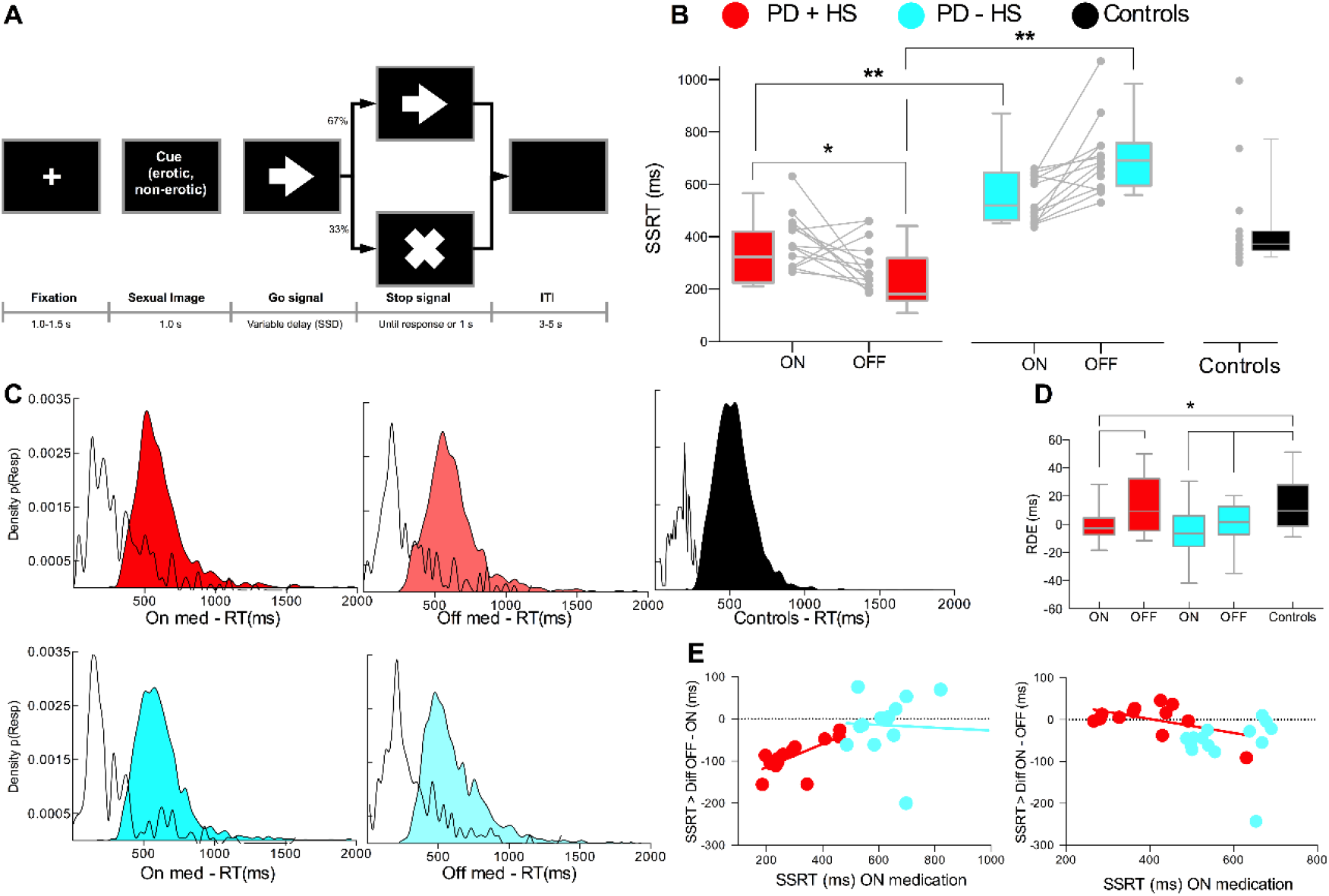
Behavioural paradigm and results. SSRT estimates as a function of *Group* and *Medication* conditions in Study 1. Study 1 **(A)** Task trial example; **(B)** SSRT for groups and individual dots per group and medication states; **(C)** Density distribution for *StopRespond* and *Go* conditions; **(D)** *Response delay effect* (*RDE*) between erotic and non-erotic go trials; **(E)** Within-subject change in *SSRT* from off to on levodopa medication is illustrated with single dots correlations showing greater benefits in response inhibition (i.e., reduction in *SSRT* from “off” to “on” status). \**p*<.05; \*\**p*<.01. Results are shown as box-and-whisker plots with each box representing the 10–90 percentile where medians are represented by internal lines.

### Erotic stop-signal task analysis

As recommended^40^, we first evaluated the independence of the go and stop processes by: a) correlations between StopRespond reaction times and correct go trials of each stimuli condition (erotic and non-erotic) per participant; b) correlations between the go trials and SSRT; and c) plotting cumulative distributions of Go and StopRespond reaction times (**Figure 1C**, **5A**).

### Behavioural, neuropsychological, and neuropsychiatric tests

A classic stop signal task was used to obtain measures of response inhibition without presence of sexual cues. General cognition and executive measures were included to discard any cognitive downsides in our sample. To this, we employed the Montreal Cognitive Assessment^41^ (as cognitive screening test), the Stroop interference test, Frontal Assessment Battery, forward and backward digit span from Wechsler Adult Intelligence Scale IV^42^), phonemic and semantic fluencies. Neuropsychiatric variables that may influence ICDs were included (Barratt Impulsiveness Scale-11^43^; Geriatric Depression Scale^44^; Starkstein Apathy Scale^45^; Beck Anxiety Scale^46^). Last, a sexual inventory test (Brief Male Sexual Function Inventory^47^) was used to include the latest sexual activities in our sample.

### Imaging data acquisition

Imaging data was collected using a 3T hybrid PET-MRI scanner (mMR Biograph, Siemens AG, Germany) with a 12-channel head array coil. Task-based fMRI was acquired using a single-shot gradient-echo echo-planar imaging (EPI) 2D pulse sequence with the following parameters: *TR/TE* = 2000/30ms; optimum flip angle using the Ernst equation i.e., 79°, *spatial resolution* = 3×3×4mm3; *field of view* = 192mm; *matrix* = 64×64; *slice thickness*= 4mm and acceleration factor of 2 (IPAT2). Five fMRI runs were acquired per session (260 volumes per run, 12mins) with 2-min pause between runs. Imaging protocol also included a 3D T1-weighted MP-RAGE (*TR/TE/TI* = 2300/3.34/900ms;*flip angle* = 8; and *isotropic spatial resolution* = 1mm3, *FoV* = 256mm, *matrix* = 256×256, *slice thickness* = 1mm); a fieldmap generated from two 2D gradient-echo images (*TR/TE1/TE2* = 455/4.92/7.38ms, *flip angle* = 60° with the spatial resolution as the fMRI EPI volumes); and diffusion weighted images (DWI) using a single-shot 2D spin-echo EPI sequence (*TR/TE* = 10000/102ms; *isotropic resolution =* 2mm). DWI were acquired for 64 non-collinear encoding directions with *b-value*=1000s/mm2 and two *b*=0s/mm2 images with opposite polarity of the phase encoding direction (AP and PA).

### fMRI preprocessing steps

T1-weighted anatomical images for all subjects were segmented into gray matter, white matter, and cerebrospinal fluid using the unified segmentation tool^48^ provided in SPM12. Gray matter maps were then transformed into the Montreal Neurological Institute (MNI) space using the DARTEL tool^49^. fMRI preprocessing was carried out with tools from FSL (FMRIB Software Library) and SPM12. Preprocessing included slice timing correction, motion correction by realigning to the first volume, correction of magnetic field inhomogeneities induced geometrical distortions and signal dropout, and co-registration to the anatomical images. Then, fMRI volumes were normalized to MNI space and smoothed with an 8mm3 FWHM Gaussian kernel and filtered over time using a high-pass filter of 128s. Omission errors and presses during sexual cues were classified as errors for the fMRI analysis.

### Task-based fMRI analysis

Task-related activities were analyzed by sexual cue types using the following regressors: Go, Stop-Inhibit, Stop-Respond, Null, and motion regressors, derivatives and quadratic forms, representing each event with an impulse function convolved with a canonical hemodynamic response function. fMRI significance was considered at *p<*.005, with FWE correction at the cluster level (*p<*.05). Small-volume correction for anatomically defined regions of interest were conducted in some regions based on a priori hypothesis.

### Dynamic causal modelling

To extract regional time series for dynamic causal modelling, we specified an F test across all trials (*F* contrast: *p*<.05) to obtain the first eigenvariate of the brain-oxygen-level dependent imaging time series from 4 volumes of interest in Stop-Inhibit erotic vs. non-erotic contrast: pre-SMA [*x*=6, *y*=24, *z*=58], ACC [*x*=0, *y*=24, *z*=36], caudate [*x*=-10, *y*=8, *z*=14] and VTA [*x*=-4, *y*=-16, *z*=-14]. Each subject’s *F* test was used to identify local maxima closest to the group peak to then extract the first eigenvariate from a 5mm sphere at the subject-specific peak.

Dynamic causal modelling^50^ estimated the effective connectivity on 13 generative models representing alternative hypotheses of the causal interactions between cortico-subcortical areas guiding hypersexuality. Structural architecture of our models (**Figure S1**) was informed by primate anatomical and human functional connectivity^51–58^. All models were constructed, fitted, and compared for each patient (**Supplementary Material**). Bayesian model selection^59^ was used to account for activity in the cortical and subcortical regions during the task and estimate the circuit abnormalities in hypersexuality^59^. Effective connectivity strength and task-modulations were compared with random-effects Bayesian Model Averaging to obtain average connectivity estimates across models and patients^59^.

### Diffusion Weighted Image Acquisition

After running whole brain diffusion tensor based tractography for each patient, the corresponding tracts from the dynamic causal modelling model were segmented. Preprocessing was performed using QSIPrep0.13.0RC1, which is based on Nipype 1.6.0^60,61^.

### Diffusion Tensor Imaging Processing

First, to ensure that diffusion tensor imaging was a viable model for this study, we inspected the different fibers directions for these tracts in the HCP1065 atlas and confirm they do not have a significant number of crossing fibers. Diffusion tensor estimation was carried out using the non-linear least-squares method implemented by Dipy. Spatial normalization in a tensor-based manner was done using dti-tk^62^. Then, tensors were normalized to a population template constructed by averaging each group of subjects. Finally, the images were mapped to 1mm^3 MNI space. Relevant features – fractional anisotropy, mean diffusivity – were extracted at this point using dti-tk. Then we ran a deterministic tracking algorithm implemented in trackvis^63^, which resulted in a whole brain tractography per subject.

For tract selection across groups, we used automated fiber quantification^64^ to compute the fractional anisotropy and mean diffusivity over the tracts, ensuring that the approximate length and shape were similar between patients. A total of 100 nodes for fractional anisotropy values were obtained for each fibre bundle. To compute the difference between groups in fractional anisotropy and mean diffusivity, we conducted t-tests over the nodes for every tract (using TFCE as statistical method) between groups followed by non-parametric permutation tests (*p<*.05, FWE Holm-Bonferroni corrections). This analysis was implemented using the code from ptsa^65^ and MNE^66^.

### Repetitive transcranial magnetic stimulation (rTMS)

In study 2, we used rTMS (Magstim Rapid2 Plus1 stimulator, Magstim, Whitland, UK) to apply intermittent theta burst stimulation^67^ over the pre-SMA [*x*=6, *y*=24, *z*=58; Stop- Inhibit erotic vs non-erotic results in Study 1; **Table 3**], consisting of trains of 10 bursts (2s) repeated every 10s for 20 cycles (600 pulses in total). Each burst consisted of a 3- pulse burst (50Hz) every 200ms with 100% intensity relative to patient’s dominant-hand flexor digitorum. Scalp-to-cortex distance correction was used for intensity adjustments^68^. A T1-weigthed MRI scan of each patient was used to determine the exact target location using Brainsight frameless stereotactic system (Rogue Research, Canada) and a Polaris infrared (NorthernDigital, Canada). Sham stimulation was performed using a sham-controlled coil (AirFilm Placebo, Magstim) controlling for device appearance and physiological properties (*i.e*., sound) of stimulation.

### Statistical analysis

Differences between conditions were analyzed using repeated-measures general linear model comparing medication (on vs. off) and condition (erotic vs. non-erotic) as the within groups variable and groups (PD+HS vs. PD-HS vs. controls) as between-group variable for successful and unsuccessful stop trials. When comparing SSRT between medication (on vs. off) and stimulation (real vs. sham), support for the alternative hypothesis (*H-:* SSRToff < SSRTon) vs the null hypothesis (*H0:* SSRToff = SSRTon) was also assessed with Bayesian paired t-tests (as implemented in JASP^69^, with default effect size priors, Cauchy scale 0.707). Results are reported as one-tailed Bayes factor (i.e., *BF_-0_* represents p(data|H-)/p(data|H0). Moreover, correlations with neuropsychological and clinical variables were executed with response inhibition measures. Brain-behaviour Pearson correlations were executed between SSRT, and beta-weights extracted from brain regions of interests, fractional anisotropy, and mean diffusivity (mean values along the segments for each tract) and with effective connectivity (dynamic causal modelling). Additional Pearson correlations between changes in SSRT after rTMS and neuropsychological variables were calculated.

### Data availability

Codes and data analysis pipelines can be found in https://github.com/cinac-cognition/tms_fmri_dti_hypersexual_pd. Applications for de-identified anonymous data can be made to HM-CINAC by contacting the corresponding author. Public access to the patient’s data is not permitted following our local ethical restrictions.

## Results

### Clinical and demographics characteristics

Groups were matched in terms of age, education, and clinical variables. While PD+HS patients also experienced other ICD subtypes in addition to hypersexuality (**Table 1**), it was the most dominant ICD subtype according to the initial interview (patient and families) and scores on the Questionnaire for Impulsive-Compulsive Disorder in Parkinson’s disease. Binge eating was the 2^nd^ most frequent subtype in our sample. Evaluation of sexual arousal before and after task performance was collected to ensure the observed behaviour and fMRI signals were based on a real sexual change (**Table 2**). A total sample size of 14 PD+HS, 14 PD-HS and 16 controls were included in the final analysis (study 1). Excluded patients were due to significant reductions in their hypersexuality between scanning sessions (*n*=2 PD+HS patients) and motion artifacts (*n*=3 PD-HS patients; *n*=1 control). Secondly, a total of 18 PD+HS patients were included in the final analysis (study 2) as reduced hypersexuality between neuromodulation sessions was seen (*n*=2). As part of the recruitment process (not included in the initial sample size), two-step interviews rejected *n*=7 patients (*n*=1 depressed patient, *n*=1 unwilling to spend time in the scanner, *n*=3 had cognitive deficits, *n*=2 homosexual patients).

**Table 2.** Sexual desire pre- vs. post-task performance and exposure to sexual visual stimuli.

|  | pre-task | post-task | *p*-values |
| --- | --- | --- | --- |
| **Off session** |  |  |  |
| PD+HS | 2.76±1.1 | 4.43±1.3 | <.001<sup>a</sup> |
| PD-HS | 0.90±0.6 | 1.10±0.7 | .18<sup>a</sup> |
| *p*-values | <.001<sup>b</sup> | <.001<sup>b</sup> |  |
| **On session** |  |  |  |
| PD+HS | 2.74±0.9 | 5.65±1.0 | <.001<sup>a</sup> |
| PD-HS | 0.60±0.5 | 0.90±0.5 | .09<sup>a</sup> |
| *p*-values | <.001<sup>b</sup> | <.001<sup>b</sup> |  |

Results obtained from average values of first two items on the arousal scale. <sup>a</sup>Paired t-test comparison; <sup>b</sup>unpaired t-test comparison.

### Behavioural results

Our ad-hoc behavioural task generated enhanced arousal in PD+HS patients while on- medication (**Table 2**). Under the race stopping model, going and stopping are supposed to act independently^70^, confirmed in our data following recommended tests^40^, such as faster StopRespond reaction times compared to go trials (**Figure 1C**; **Supplementary Material**). Group and medication effects modulated differently response inhibition [**Table S1**; **Figure 1B**; medication x group, *F*(2,44)=5.92, *p*=.006; group, *F*(2,44)=7.07, *p*=.002], where post-hoc tests showed significant medication effects in PD+HS [*t*(12)=- 3.06, *p*=.01] with strong evidence that response inhibition was impaired by medication and sexual cues [*BF_-0_*= 11.60; see sequential analysis in **Figure S2**]. Performance before medication (off medication) explained improved SSRT in PD+HS (positive correlations between SSRT on-off medication difference and on medication; *r*=.753, *p*=.003; **Figure 1E**). Proactive response delay effects were modulated by group [*F*(1,44)=4.11, *p*=.02], explained by increased response adaptation in controls compared to PD+HS (on medication, p=.02) and PD-HS patients (on medication, *p*=.01; **Figure 1D**) and by medication [*F*(1,44)=4.52, *p*=.04], as evident from faster adaptation while on medication in PD+HS [*p*<.03; **Figure 1D**]. As expected, classic measures of response inhibition (without erotic cues) revealed no impairments in PD+HS compared to controls (**Supplementary Material**).

### fMRI results

#### Neural Network for inhibition under erotic influence

To decipher the key areas behind successful inhibition (Stop-Inhibit) under sexual influence, a general linear model on group, condition and medication revealed a medication by group interaction and medication effect over superior parietal lobe activity (**Table 3**; **Figure 2A**). This effect was driven by the PD+HS group also significantly recruiting the ACC, middle frontal gyrus, caudate and VTA (**Table 3**; **Figure 2B**). Meanwhile, a group effect showed increased activity over the pre-SMA in the PD+HS compared to PD-HS group (on medication; **Table 3**; **Figure 2C**). Moreover, successful inhibition of actions (compared to unsuccessful inhibition) revealed a main effect of medication over superior parietal lobe, ACC, and supplementary motor complex (**Table S3**; **Figure S3A**). This effect was mediated by the PD+HS group (on vs off medication) showing enhanced activity in frontal pole, supplementary motor area and inferior frontal gyrus (**Table S3**; **Figure S3B**).

**Table 3.**
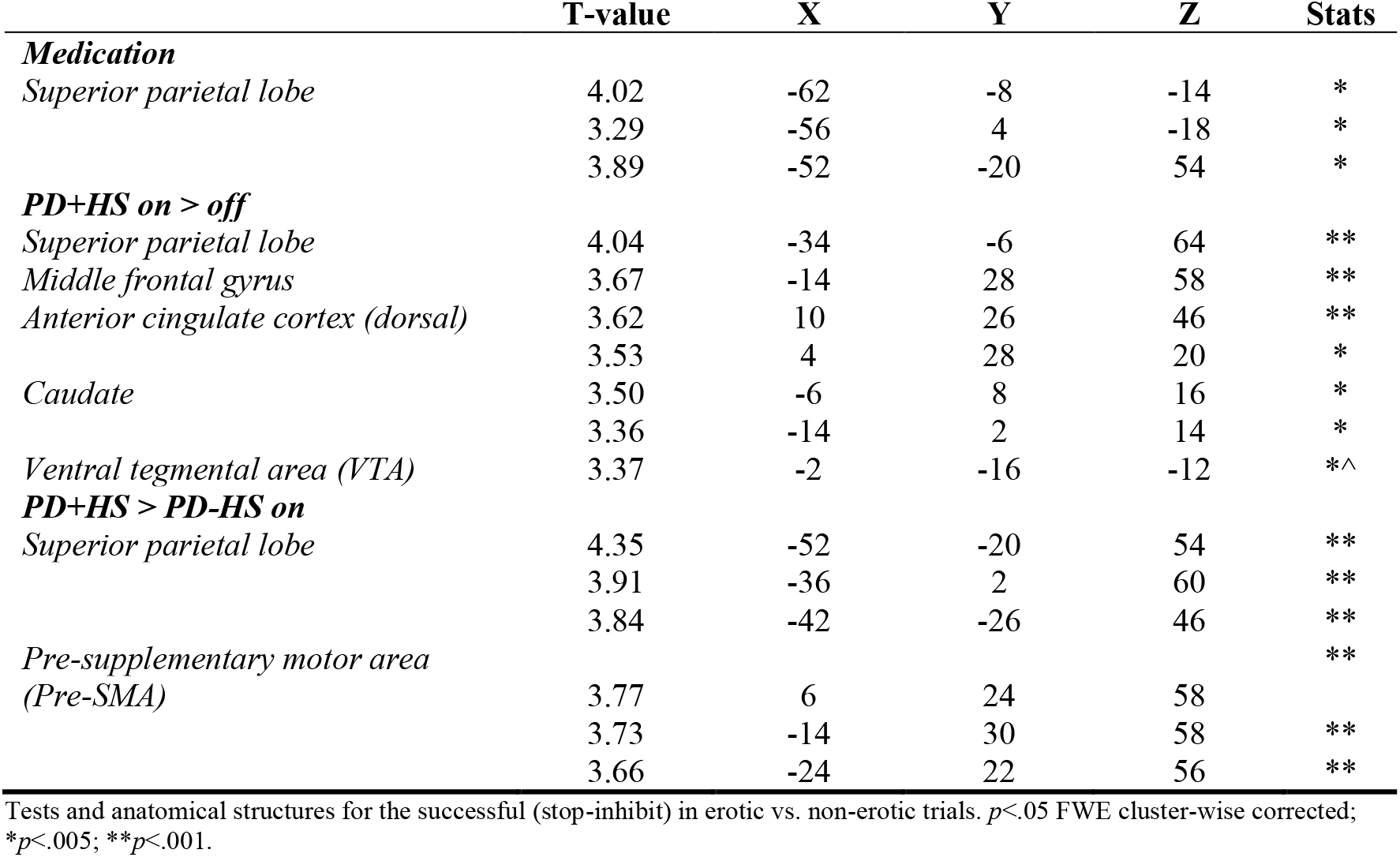
Stop-Inhibit in Erotic vs. Non-Erotic trials.

**Figure 2.**
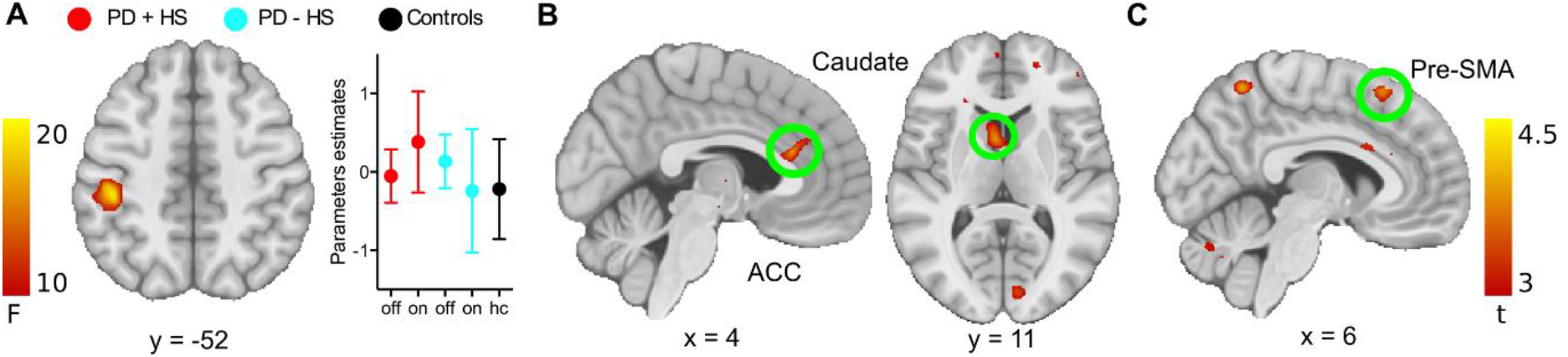
Neural correlates for successful inhibition in erotic vs non-erotic conditions. **(A)** *Medication* x *Group* interaction with parameter estimates over superior parietal cortex per group; **(B)** *Medication* effect revealing increased activity in PD+HS comparing on vs. off medication states; **(C)** *Group* effect revealing enhanced activity in the PD+HS group compared to PD-HS (on medication).

#### Neural correlates of successful and unsuccessful inhibition under erotic influence

Following sexual cues, successful (Stop-Inhibit) compared to unsuccessful inhibition showed a disease x medication interaction as well as in medication effects in the postcentral gyrus over the parietal cortex (**Table S4**). Post-hoc tests showed significant activity increase in the PD+HS group (compared to PD-HS group while medicated) in the precuneus and supplementary motor area as well as the superior parietal lobe (**Table S4**). In contrast, when looking at the specific activity patterns in unsuccessful inhibition for the erotic (against non-erotic trials), we observed a disease x medication interaction over the ACC (**Table S5**). This activity was guided by the PD-HS group, which showed significantly higher activity than PD+HS patients in the ACC while medicated (**Table S5**).

#### Unsuccessful inhibition under erotic influence

Given hypersexuality may be driven by failure to control ongoing behaviour when exposed to sexual cues, we analysed the erotic influence over failed inhibition at the neural level. Failing to stop specifically under erotic influence (erotic vs. non-erotic trials) revealed a significant medication x group interaction over the superior parietal lobe, supplementary motor area and superior frontal gyrus (**Table S5**). Such interaction revealed in PD+HS patients (on vs. off) a significant cluster activity over the left caudate (*p*<.005, small volume correction FWE corrected; **Table S5**), a region that is also present during Stop-Inhibit in erotic trials.

#### Cortico-subcortical influences underlying response inhibition

The cortico-subcortical model comparison revealed striking results: there was strong evidence in favour of one model above all others (model 12; **Figure 3B**). Across groups, the pre-SMA to both caudate and ACC connections were excitatory (**Figure 3A**). Importantly, significant differences in pre-SMA-caudate connectivity were seen between PD+HS (0.004±0.006) and PD-HS patients (0.02±0.03) while medicated (*p*=.02; **Figure 3C**). Moreover, pre-SMA-caudate connectivity was modified by medication in PD+HS (on vs. off: *p*=.03; **Figure 3C**). Local intrinsic connectivity in the caudate correlated with SSRT in PD+HS patients while medicated (*r*=.55, *p*=.04), which reflects local neuronal adaptation, suggesting an altered neurobiological link to response inhibition changes.

**Figure 3.**
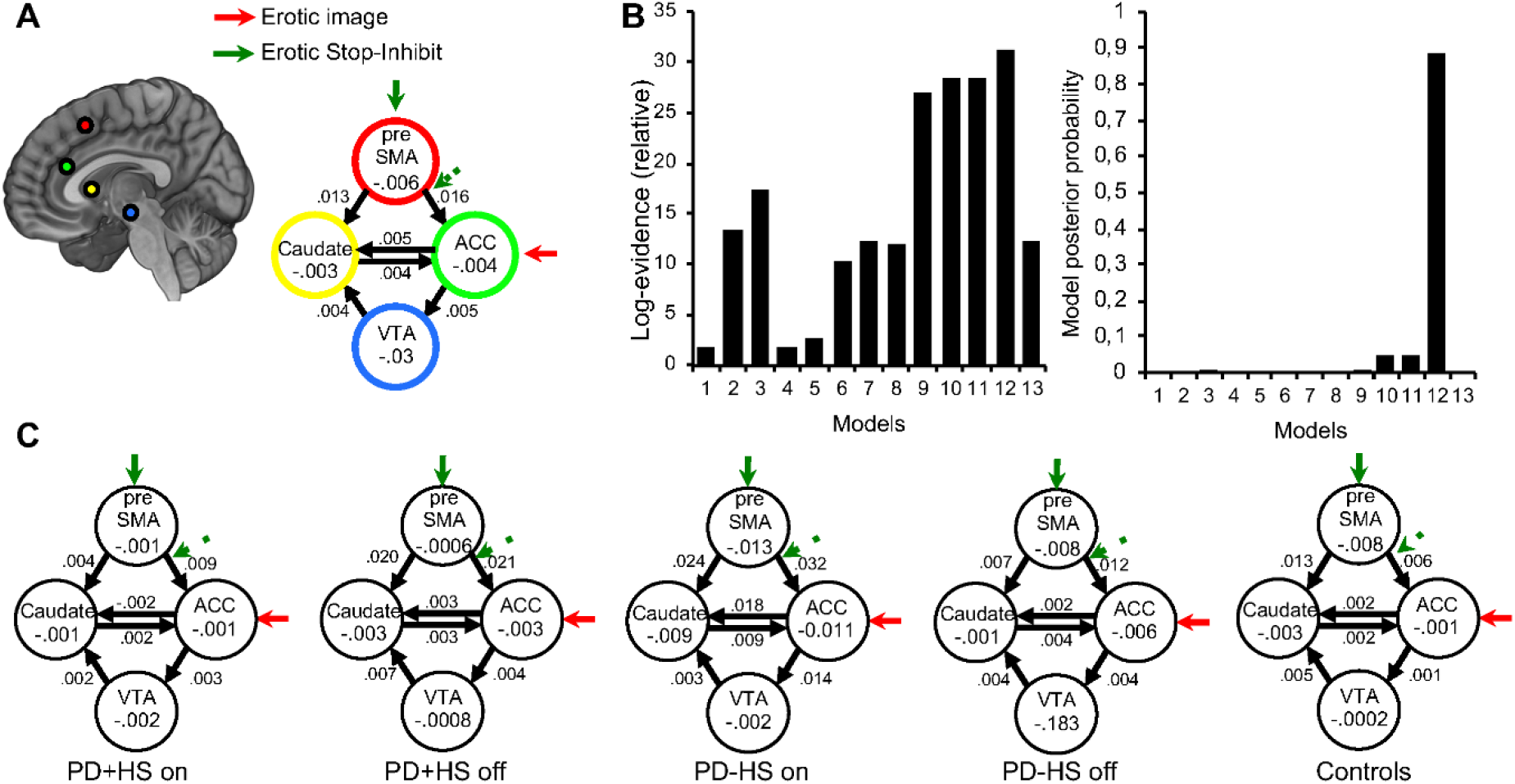
Dynamic causal modelling (DCM), winning model and group differences. **(A)** Bayesian Model Selection (BMS): dotted arrows indicate modulatory effects of stopping (correct stop-inhibit) and erotic cues. Solid bold arrows indicate driving inputs of task performance locally. Models share local effects of task on pre-SMA structure. The most likely model counts with increased excitatory connectivity from the pre-SMA to the caudate and ACC (DCM.A average connectivity). This connectivity is modulated locally in pre-SMA by task (DCM.B) and by modulatory inputs in descending connection towards ACC (DCM.A). Bidirectional connections between ACC and caudate are detected (DCM.A); **(B)** Using the free-energy estimate of the log-model evidence between models, very strong evidence was found in favor of one model (Figure 3A). The group posterior probability (close to 1) illustrates the strong support for the most likely model; **(C)** Model connectivity values after Bayesian parameter averaging at the group level reveal reduced excitatory connectivity in pre-SMA to caudate pathway between PD+HS and PD-HS patients. Ultimately, local task modulation over pre-SMA differed between patients.

#### Local modulatory effects of Stop-Inhibit

The winning effective connectivity model revealed a task modulation (Stop-inhibit in erotic vs non-erotic) over pre-SMA, significantly different between PD+HS (0.04±0.11) and PD-HS groups (-.10±0.16; *p*=.01; **Figure 3C**). This finding is indicative of local pre-SMA differential responses to stop cues between patients may contribute descending aberrant signals to control behaviour.

#### Structural connectivity

We observed bilateral significant differences in the pre-SMA-caudate tract, showing higher fractional anisotropy values and lower mean diffusivity (**Figure 4B**) in PD+HS compared to PD-HS group and controls (**Figure S4**). Interestingly, the opposite pattern is seen for the caudate-ACC tract with lower fractional anisotropy but also reduced mean diffusivity values in PD+HS compared to PD-HS patients; **Figure 4C**). The VTA-caudate tract could not be robustly reconstructed with the diffusion tensor imaging model and the proposed tractography algorithm, thus excluded from the analysis. Along the right pre-SMA-caudate tract, mean diffusivity values showed a positive SSRT correlation for PD- HS patients (*r*=.79, *p*=.01; **Figure S5**) but a non-significant negative correlation in PD+HS (off medicated, *r*=-.20; *p*=.61; **Figure S5**). Thus, structural connectivity predicted behavioural changes between patients’ groups.

**Figure 4.**
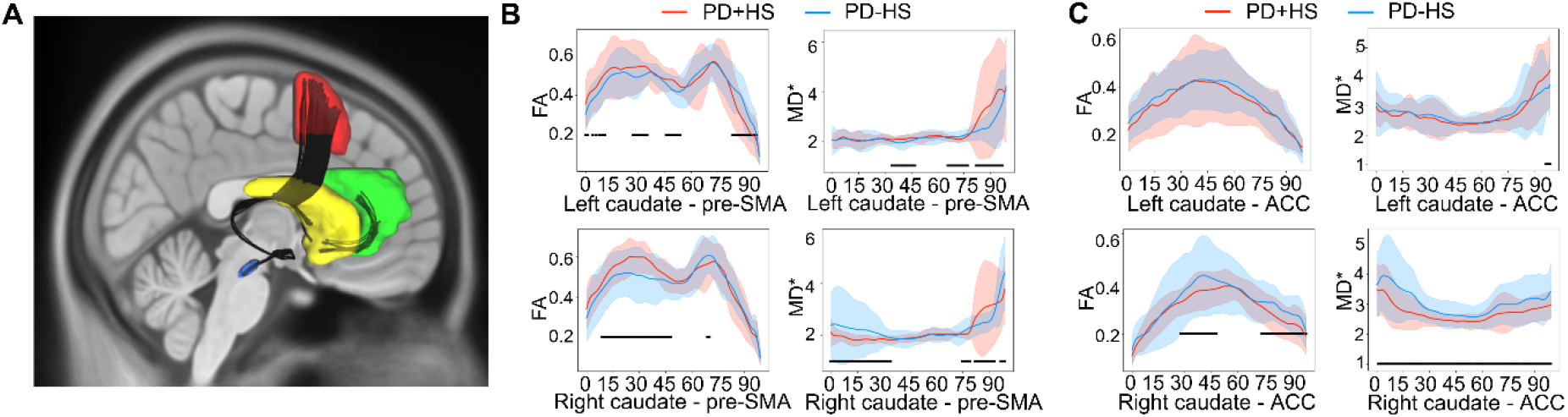
Tracts representation and diffusion tensor imaging (DTI) scalar measures – fractional anisotropy (*FA*), mean diffusivity (*MD*\*). Comparisons included PD+HS (*n*=10) and PD-HS patients (*n*=14) and controls (*n*=13); **(A)** Tracts segmentation between areas in the dynamic causal modelling (DCM; VTA, caudate, ACC, pre-SMA) in the HCP1065 template, superposed on a T1 MNI template; (B); (C) *FA* and *MD* along the segments of each tract. Significant differences (*p*<.05 corrected) along the segments between the groups are underlined in black (see Supplementary Material for more details about the thresholding procedure).

#### Structural connectivity predicts effective connectivity

We tested fractional anisotropy and mean diffusivity correlations with dynamic causal modelling parameters in cortico-subcortical tracts. A relevant approaching significance negative correlation was found between fractional anisotropy and dynamic causal modelling from the right pre-SMA and caudate connection in PD+HS patients (*r*=-.69, *p*=.06). Thus, higher fractional anisotropy in PD+HS patients corresponds to lower effective connectivity (**Figure S6**), consistent with pathological compensatory responses to preserve functional connectivity^71,72^.

#### Non-invasive brain stimulation: repairing hypersexual behaviour

Following real intermittent theta burst stimulation over the pre-SMA (compared to sham), significant improvement of response inhibition was found in PD+HS patients for erotic (*p*=.02) and non-erotic (*p*=.04) stimuli (**Figure 5B**), an effect showing moderate evidence towards the alternative hypothesis [*BF-_0_*=3.40; sequential analysis in **Figure S7**]. Moreover, faster Go reaction times in both erotic (*p*=.06) and non-erotic (*p*=.04) stimuli were seen across patients (**Figure 5C**). Independence race-model assumptions were also confirmed in this study (**Figure 5A**; **Supplementary Material**). Correlation analysis between response inhibition and backwards digit span correlated with larger improvements in SSRT after intermittent theta burst stimulation (*r*=.49; *p*=.03). Finally, results were not affected by possible session order effects (**Supplementary Material**).

**Figure 5.**
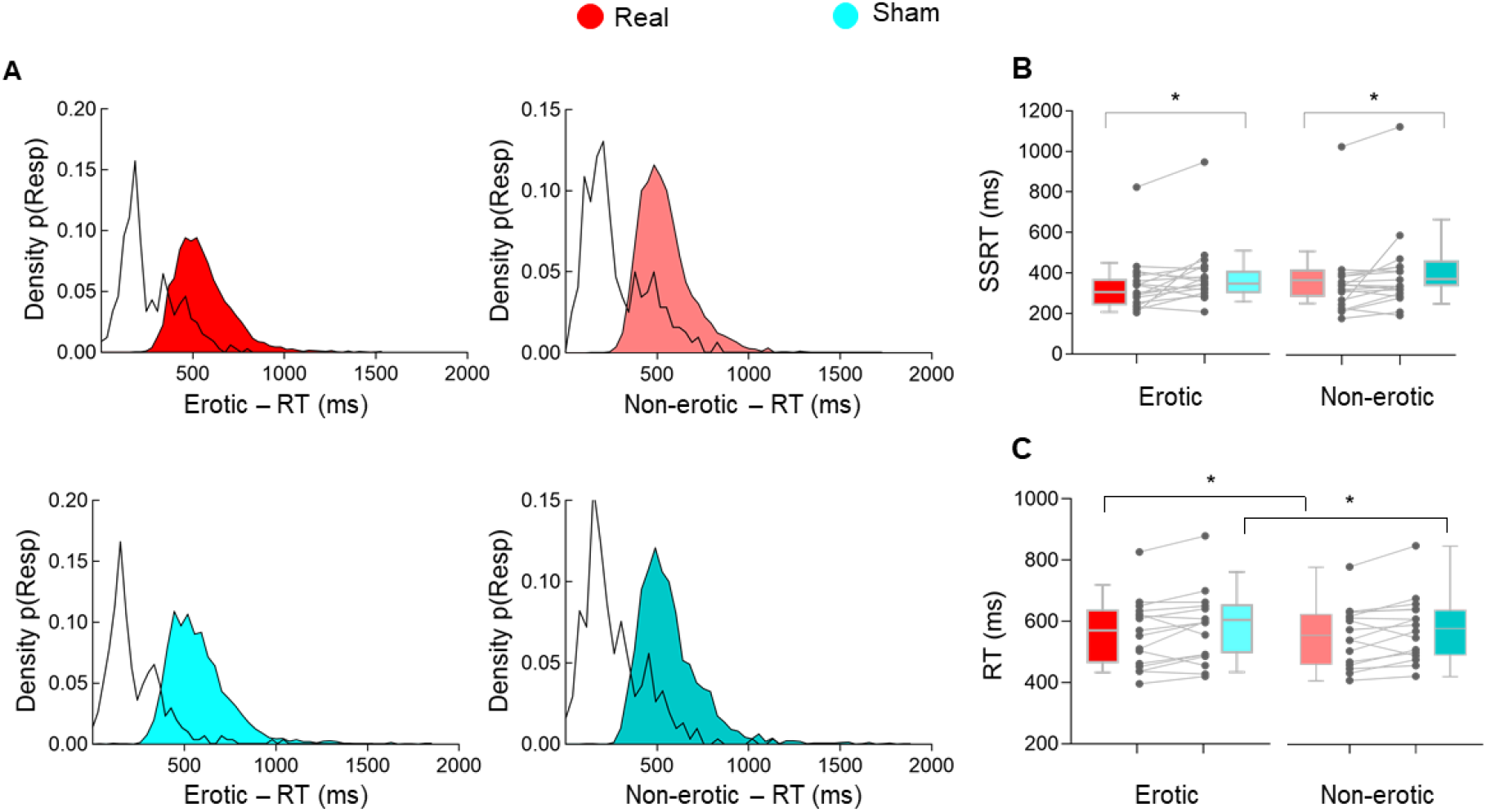
Neuromodulation effects on cognitive control and hypersexuality. Study 2. **(A)** Density distribution for *StopRespond* (white distribution) and *Go* trials (coloured) per stimulation session and per trial type; **(B)** Boxplot and individual values for *SSRT* and **(C)** *Go* trials reaction times for erotic and non-erotic trials after real and placebo stimulations. \**p*<.05; Results are shown as box-and-whisker plots with each box representing the 10–90 percentile where medians are represented by internal lines.

## Discussion

Impaired response inhibition was found in PD+HS when exposed to sexual cues while on medication. The cortico-striatal regions responsible for uncontrolled sexual behaviours included inhibitory hubs such as the pre-SMA, but also the VTA, caudate and ACC as part of the limbic system. Effective connectivity models revealed less engagement of pre-SMA in stop-related trials and reduced functional connectivity with the caudate in PD+HS compared to PD-HS patients, showing opposite SSRT correlations between patients. This functional change was held by enhanced fractional anisotropy and reduced mean diffusivity in the pre-SMA-caudate pathway in PD+HS. Importantly, excitatory neuromodulation over the pre-SMA improved response inhibition under sexual influence in PD+HS patients. Our results provide compelling evidence to understand the neurobiological underpinnings of hypersexual behaviour and reveal, to the best of our knowledge, a first attempt using non-invasive neuromodulation to repair hypersexuality in Parkinson’s disease.

### Sexual cues impair response inhibition in PD+HS patients

We observed greater disruption of response inhibition upon presence of sexual cues driven by dopaminergic medication (**Figure 1B**). Moreover, proactive control between conditions was reduced while on medication (**Figure 1D**), suggesting some form of anticipatory maladaptation in PD+HS. In other words, dopaminergic drugs impaired controlling systems needed to respond upon sexual signals, since PD+HS patients showed no impairments in response inhibition in a classic stop signal task. Hence, our results add notions of updated behavioural methods (ad-hoc stimuli related to the clinical problem) to test cognitive disturbances and help solving current debates on why some studies report unimpaired response inhibition in ICD^27,28^.

Given reward sensitivity increases in Parkinson’s disease patients while medicated^73,74^, our results support a bias towards enhanced incentive salience that puts constraints on the controlling functions of the addicted brain. The incentive salience hypothesis^75^ suggests combinations of previously acquired cue–reward associations may put significant constraints over behaviour. In addition, the impaired response inhibition and salience attribution model proposes both systems as responsible in guiding patients towards addiction^76^. Given emotional inputs deactivate essential cognitive control regions^77^, presence of previously experienced sexual cues shall downgrade top-down control and permit uncontrolled sexual behaviour to occur. Thus, prior reports showing intact cognitive control abilities in ICD^27,28^ may be re-framed as a net effect on inhibition, as no inputs from cue–reward associations were currently influencing patients’ behaviour. Our results go in line with both enhanced sensitivity to sexual cues together with reductions in response inhibition ability, consistent with the inhibition and salience attribution model^76^.

### Dysfunctional circuitry in hypersexuality: functional and anatomical changes

The circuitry mediating sexual events in healthy people activates limbic areas such as the ventral striatum, amygdala, ACC, parietal and orbitofrontal cortex^78–81^. Controlling sexual actions, in contrast, recruits specific regions of the inhibitory network including the pre-SMA, inferior-frontal gyrus, superior parietal cortex and ACC^82–84^. Here, we found increased limbic (including ACC, VTA and caudate) and pre-SMA activity during response inhibition under sexual influence in PD+HS patients (on vs. off medication) compared to PD-HS patients (**Figure 2C**).

We interpret these findings as an abnormal mesocorticolimbic enhancement without controlling functions leading to PD+HS. Upon a faulty mesocorticolimbic circuitry due to dopamine agonists, including the VTA, ACC and ventral striatum^85,86^, enhanced “wanting” may thus lead to “hard to resist” reward compulsive search^38,87,88^. Hence, the combined “wanting” and “hard to resist” elements may integrate the neurobiological foundations of hypersexual behaviour often found in Parkinson’s disease patients. PD+HS patients reported heightened sexual desire after task performance (**Table 2**), which may require a larger cortical recruitment to generate some form of controlled behaviour. Thus, a combination of dysfunctional dopaminergic bottom-up (limbic) and top-down (executive) control under medication may boost uncontrolled desire in our patients, explained by the combination of regions here reported. This view matches well according to the dual control model of sexual response^89^, where a shared duality between sexual excitatory and inhibitory mechanisms regulates sexual behaviour. Together with their model, plasticity alterations after persistent reward cue-exposure^75,90^ may predispose one of the two dual systems to set disproportional high sexual excitation or disproportional low sexual inhibition. Our results support the view that an enhanced sexual excitation system along the mesocorticolimbic system finds no or reduced opposition from prefrontal top-down control regions, leading to perturbed control of sexual behaviour.

Yet, it is difficult to understand how the causality and directionality of active task-based fMRI regions are involved in PD+HS. As revealed by a winning functional connectivity model (**Figure 3B**), the pre-SMA and its connectivity with the caudate put forward 2 pathophysiological assumptions in the hypersexual circuit: the pre-SMA showed a reduced response to task-related signals and weakened its excitatory connection with the caudate (**Figure 3C**). Such top-down dysfunctional connection (based on dynamic causal modelling) was linked to an anatomical increase (based on fractional anisotropy tracts) in PD+HS (approaching significance values). Ultimately, opposite correlations between patients were seen between the pre-SMA-caudate tract and SSRT, which altogether suggests a dual neurobiological problem (local and descending pre-SMA connectivity) in the cognitive control network^91,92^. Thus, reductions in top-down control rely upon paradoxical increases of white matter integrity, a possible neural compensatory response previously reported in Parkinson’s disease and ICD^72,93,94^.

The neurobiological changes in pre-SMA to caudate connectivity in our PD+HS sample may be explained by repeated exposure to sexual cues and plasticity changes, boosted by dopaminergic medication. Compulsive overuse of sexual acts may lead to frequent engagement and overflow activity in pre-SMA-caudate connections, which could re- organize the local activity and white matter pathways^72^. Possibly, aberrant plasticity along this pathway may increase the vulnerability to develop PD+HS and other forms of addictive behaviours as well. Together with notions that the mesocorticolimbic circuitry is overstimulated in Parkinson’s disease and ICD^4^, pre-SMA-caudate tract may be a susceptible pathway to drug-induced dopaminergic overstimulation and hypersexuality in Parkinson’s disease. Hence, considering the well-known plasticity changes associated to reward cue-exposure^90^, our results are interpreted by a remodeling framework of inhibitory brain dynamics, possibly mediated by experience-dependent synaptic plasticity. This view would explain why some Parkinson’s disease patients develop a particular form of ICD, coherent with founded notions of personality traits that influence generation of ICD in Parkinson’s disease^5,95^. Yet, further information is needed to determine whether these neurobiological changes are causal or putative consequences of dopaminergic medication in PD+HS.

### A route towards rehabilitation: neuromodulation to repair hypersexuality

To date, hypersexuality counts with no direct validated treatment. With aim improving this deficient clinical scenario, we compared excitatory rTMS stimulation over the pre-SMA to a sham condition and found an acute benefit on response inhibition under sexual influence in PD+HS patients (**Figure 5B**). We favored stimulation of the pre-SMA based on 2 foundations: its dominant role in response inhibition^20,23^ and our fMRI group-based findings showing pre-SMA causal downregulation in stop-related trials in PD+HS. Based on previous rTMS beneficial effects over response inhibition^96–98^ and the excitability properties of intermittent theta burst stimulation over the pre-SMA while performing inhibitory tasks^98^, we assume our protocol enhanced pre-SMA inhibitory function. Similarly, changes in the connectivity between pre-SMA and caudate may explain the behavioural enhancement due to their direct functional connection in response inhibition tasks^91^. Thus, intermittent theta burst stimulation may reset dysfunctional pre-SMA downstream pathways and induce distant control over limbic networks in PD+HS patients.

An alternative explanation of our neuromodulation findings is a possible variation on the emotional regulation associated to sexual cues. Previously, a single-blind, randomized crossover study investigated the impact of pre-SMA stimulation (online interference with 5 pulses at 10Hz; sham-controlled) on a facial emotion recognition task^99^. After stimulation periods, a specific downregulation in happiness recognition was disrupted compared to sham^99^, suggestive of an influence on the rewarding system when stimulating the pre-SMA. In line with such findings, our protocol could have produced an impact on how sexual cues were identified in our patients and thus may have placed less constraints over response inhibition.

Recent evidence suggests the efficacy of rTMS in improving response inhibition in Parkinson’s disease when applied over the dorsolateral prefrontal cortex^100^. Yet, to our best knowledge, no studies have targeted the pre-SMA with aim modifying cognitive control in Parkinson’s disease with or without ICDs. Interestingly, however, pre-SMA stimulation (1Hz) improved Parkinson’s disease-related dyskinesias^101^ – involuntary movements induced by dopaminergic mediation – that might belong to a related continuum of the motor circuits. Hence, their results may parallel our findings as their stimulation protocol may have temporarily modified related pathophysiological mechanisms^4^ as those speculated here and possibly may offer a common target to lessen dopaminergic side-effects.

### Strengths, limitations, and conclusions

One strength of this study was using a highly selected clinical sample and ad-hoc stimuli to provoke PD+HS measured with multi-level neuroimaging tools. We then used a circuit-based approach to stimulate a putative brain target essential in PD+HS. Our findings are based on a moderate sample size that should be expanded in larger cohorts. Reasons for this sample size are directly related to the scientific question that requires to limit recruitment to a precise clinical characteristic (with added recruitment efforts). Yet, both studies follow adequate sample sizes based on previous neuroimaging^10,38^ and rTMS studies on Parkinson’s disease and ICD samples^35^. Moreover, our Bayesian sequential analysis suggests the accumulation of evidence in favor of the alternative hypothesis increases relatively steep as subjects are tested toward strong and moderate evidence in study 1 and 2, respectively.

Stimulation of pre-SMA was conducted in PD+HS patients and not patients without the behavioural problem. Thus, this design constraint renders our results not specific to hypersexual issues. Yet, using sham-controlled stimulation (with ad-hoc sham rTMS equipment) provides specificity of the neuromodulation target and protocol. Moreover, given most Parkinson’s disease patients without ICD have no associated non-motor symptoms directly associated with pre-SMA functions (executive functions, response inhibition, conflict resolution, etc.), ethical issues are applicable when testing stimulation protocols in patients without a primary motive.

## Conclusion

Evidence for an incentive sensitization framework is provided to explain the interplay between desire enhancement and response inhibition in hypersexuality. We reveal a circuit-based route that deciphers the cortico-striatal anatomical and functional dysfunction and a candidate brain target for neuromodulation to change behavioural expression in patients with PD+HS.

## Data Availability

All data produced in the present work are contained in the manuscript

## Supplementary material

Supplementary material is available.

## Acknowledgements

We are thankful for patients and their families for active engagement in the studies. We also are grateful to Federación de Parkinson for helping with advertising the study.

## Methods

### The erotic stop-signal task

Following the recommendations for using the stop signal paradigm^1^, we first evaluated the independence of the go and stop processes by: a) correlations between StopRespond reaction times (RTs) and correct go trials of each stimuli condition (erotic and non-erotic) per participant; b) correlations between the go trials and stop signal reaction time (SSRT); and c) plotting cumulative distributions of Go and StopRespond RTs (**Figure 1C, 5A**). Bonferroni correction for multiple comparisons were used in *post-hoc* tests and specified when tests did not survive corrections.

### Dynamic causal modelling (DCM)

DCM estimates the effective connectivity between brain regions according to (i) average connections between regions (DCM.A), (ii) modulatory task influence on connections (erotic images and Stop-Inhibit in erotic trials, see **Figure 3A**; DCM.B) and (iii) condition-specific inputs that drive network activity (namely regional engagement in a Stop-Inhibit task; DCM.C). Across all 13 models, driving inputs (DCM.C) represented 2 key elements of the task: erotic images and Stop-Inhibit tasks. This was set to both ACC and pre-SMA areas (**Figure S1**). Bayesian parameter averaging and was used to estimate the connectivity values of the most likely model at the group level^2^.

### DWI acquisition and preprocessing steps

Marchenko-Pastur distribution principal component analysis (MP-PCA) denoising following MRtrix3’s dwidenoise^3^ was applied with a 5-voxel window. After MP-PCA, B1 field inhomogeneity was corrected using dwibiascorrect from MRtrix3 with the N4 algorithm^4^. After B1 bias correction, the mean intensity of the DWI series was adjusted so all the mean intensity of the *b*=0 images matched across each separate DWI scanning sequence. FSL (version 6.0.3:b862cdd5) eddy was used for head motion correction and Eddy current correction^5^. Eddy was configured with a *q*-space smoothing factor of 10, a total of 5 iterations, and 1000 voxels used to estimate hyperparameters. A linear first level model and a linear second level model were used to characterize Eddy current-related spatial distortion. *q*-space coordinates were forcefully assigned to shells. Field offset was attempted to be separated from subject movement. Shells were aligned post-eddy. Eddy’s outlier replacement was run^6^. Data were grouped by slice, only including values from slices determined to contain at least 250 intracerebral voxels. Groups deviating by more than 4 standard deviations from the prediction had their data replaced with imputed values. *b*=0 reference image was collected with reversed phase-encode blips, resulting in a pair of images with distortions going in opposite directions. Here, *b*=0 images with reversed phase encoding directions were used along with an equal number of *b*=0 images extracted from the DWI scans. From these pairs the susceptibility-induced off-resonance field was estimated^7^. The fieldmaps were ultimately incorporated into the Eddy current and head motion correction interpolation. Final interpolation was performed using the jac method.

Several confounding time-series were calculated based on the preprocessed DWI: framewise displacement (FD) using the implementation in *Nipype* (following the definitions by Power *et al.*^8^) The head-motion estimates calculated in the correction step were also placed within the corresponding confounds file. Slice-wise cross correlation was also calculated. The DWI time-series were resampled to ACPC, generating a preprocessed DWI run in ACPC space with 1.2mm isotropic voxels. Several internal operations of *QSIPrep* use *Nilearn* 0.7.0^9^ (RRID:SCR_001362) and *Dipy*^10^.

### Tract selection

We ran the default fiber tracking algorithm using DsiStudio^11^ with 1000000 seeds to select the tracts of interest (HCP1065 atlas). Based on the DCM results, we selected ROIs extracted from the AAL_2mm atlas (except for VTA: AAL_1mm) and the pre-SMA (HMAT 2mm atlas)^12^. Tracts were obtained by setting the first region as seed (e.g., caudate) and the end region as end (e.g., pre-SMA). However, the right caudate to ACC tract were both set as ROI to obtain a higher number of streamlines.

Then, streamlines were cut to avoid limiting access into the seed region. Finally, we deleted residual short tracts produced by cutting the tracts outside the seed region by setting the threshold to 10mm. For left caudate-ACC, we additionally deleted repeated tracts using the corresponding DsiStudio function (*voxel threshold* = 1). For right VTA- caudate, the straight streamline was removed since the tracking algorithm was not able to recognize the tracts streamlines from one of the hemispheres.

These bundles were used as a reference to extract the corresponding bundles from each subject. For this, we set to 20 the number of points to define each streamline and then we ran the recoBundles method implemented by Dipy with the default options.

## Results

### Behavioural results

While *p*(inhib) did not significantly differ between groups [*F*’s>1], it varied from 50% (**Table S1**) thus using the integration SSRT method is recommended^1^. Moreover, probability distributions were faster for StopRespond RT compared to go trials in every group [Kolmogorov-Smirnov: *p*’s>.05; except for PD-HS off medication *p*=.06; **Figure 1C**]. Last, independence between going and stopping on erotic trials was confirmed with non-significant correlations between both measures (go and SSRT) across groups [*p*’s>.05, *r*’s>.4]. The faster StopRespond RT compared to Go RT independence between go and stop are all in line with a race model of action cancellation. Mean stop signal delay (SSD) across groups did not vary significantly between groups nor between conditions (**Table S1**).

Response initiation assessed by Go trials revealed an effect of condition [*F*_(2,44)_=7.59, *p*=.009] and condition x group interaction [*F*_(2,44)_=4.45, *p*=.017] were found. Post-hoc comparisons on Go RTs did not survive Bonferroni corrections. The length of SSD was not different across groups, nor it revealed significant interactions [*F*’s>1].

Sexual influence over response inhibition (SSRT) was highly dependent on medication, group and condition (group [*F*_(2,44)_=7.07, *p*=.002]; medication x group [*F*_(2,44)_=5.92, *p* =.006]; medication x condition [*F*_(2,40)_=8.25, *p*=.006]; medication x group x condition [*F*_(2,40)_=5.96, *p*=.005]). Specifically, faster SSRT in erotic trials were seen in PD+HS compared to PD-HS both in on [*t*_(27)_=-3.25, *p*=.003] and off medications [**Table S1**, *t*_(27)_=- 8.72, *p*<.001]. Similar differences were present between patients groups in non-erotic conditions while on [*t*_(27)_=-3.72, *p*=.001] and off medications [**Table S1**, *t*_(27)_ =-6.57, *p*<.001]. The group effects were marked by faster SSRT (erotic condition) in PD+HS (off medicated) compared to PD-HS [*on*: *t*_(27)_=-3.25, *p* =.003; *off*: *t*_(27)_ =-8.72, *p*<.001] and controls [**Table S1**, *t*_(28)_=-2.38, *p*=.02]. In contrast, PD-HS showed slower inhibition in both erotic and non-erotic conditions compared to controls [**Table S1**, *t’s*>2.1; *p’s*<.3]. Medication effects across conditions revealed an specific worsening of inhibition in PD+HS patients while on compared to off medication [**Table S1**, *t*_(12)_=-3.06, *p*=.01], an effect not seen in the non-erotic condition [**Table S1**, *t’s*<.1; *p*<.8]. Error rates were not significantly different across groups, medication, or condition [*z*’s<1]. Significant difference in response adaptation on the response delay effect (RDE) was seen (Go erotic minus Go non-erotic RTs) in the PD+HS group between medication states [*on*: -.22±11.9; *off*: 13.87±17.48; *p*<.03], while non-significant in the PD-HS group [*on*: -5.40±2.3; *off*: 0.84±14.44; *p*<.35].

### Classic stop signal task

To determine whether impulsive actions were associated only to presence of erotic stimuli, we assessed performance on a classic stop signal task (without presence of erotic images). Patients and controls probabilities of inhibition were not close to 50% although no significant differences were found between groups [*F*_(2,44)_=.31, *p*=.73]. The SSRT showed significant differences between groups [*F*_(2,44)_=7.13, *p*=.002], whereby post-hoc tests result in prolonged SSRT in the PD-HS group compared to PD+HS patients [*t*_(28)_=- 3.07, *p*=.005] and controls [*t*_(28)_=3.07, *p*=.001]. The remaining variables on the classic stop signal task showed non-significant differences between the groups.

### fMRI results

#### Neural Network for successful inhibition under erotic influence

To eliminate confounding factors of the motor response embedded within inhibition of ongoing actions, Stop-Inhibit vs. Go trials were compared showing a main Group effect, with significant clusters over the motor cortex and cerebellum (**Table S2**). Such effect was driven by the PD-HS group showing greater ACC activity than PD+HS hypersexual patients (off medication; **Table S2**).

### Non-invasive brain stimulation: aiming to restore hypersexual behaviour

Probability of inhibition substantially deviated from 50% in both real and sham (∼70%; **Table S5**), thus we used the integrative approach to estimate SSRT values No significant differences in SSD were observed in any type of stimuli after either condition (**Table S5**).

Although not statistically significant, after real intermittent theta burst stimulation (compared to sham) patients produced less premature errors – anticipations during the sexual cue– in both erotic and non-erotic stimuli and tended to produce faster RTs in go trials (**Table S5**). Analyses were repeated comparing first vs. second sessions (independently of stimulation condition) to control for potential undesired order effects. No significant order effect was observed for any of the task variables including SSRT [*<u>F</u>*’s>1]. No significant difference in response adaptation on the response delay effect (*RDE*: Go erotic minus Go non-erotic RTs) was seen in any condition [*real*: 12.01±21.08; *sham*: 9.42±17.48; *p*=.90].

---

**Figure S1.**
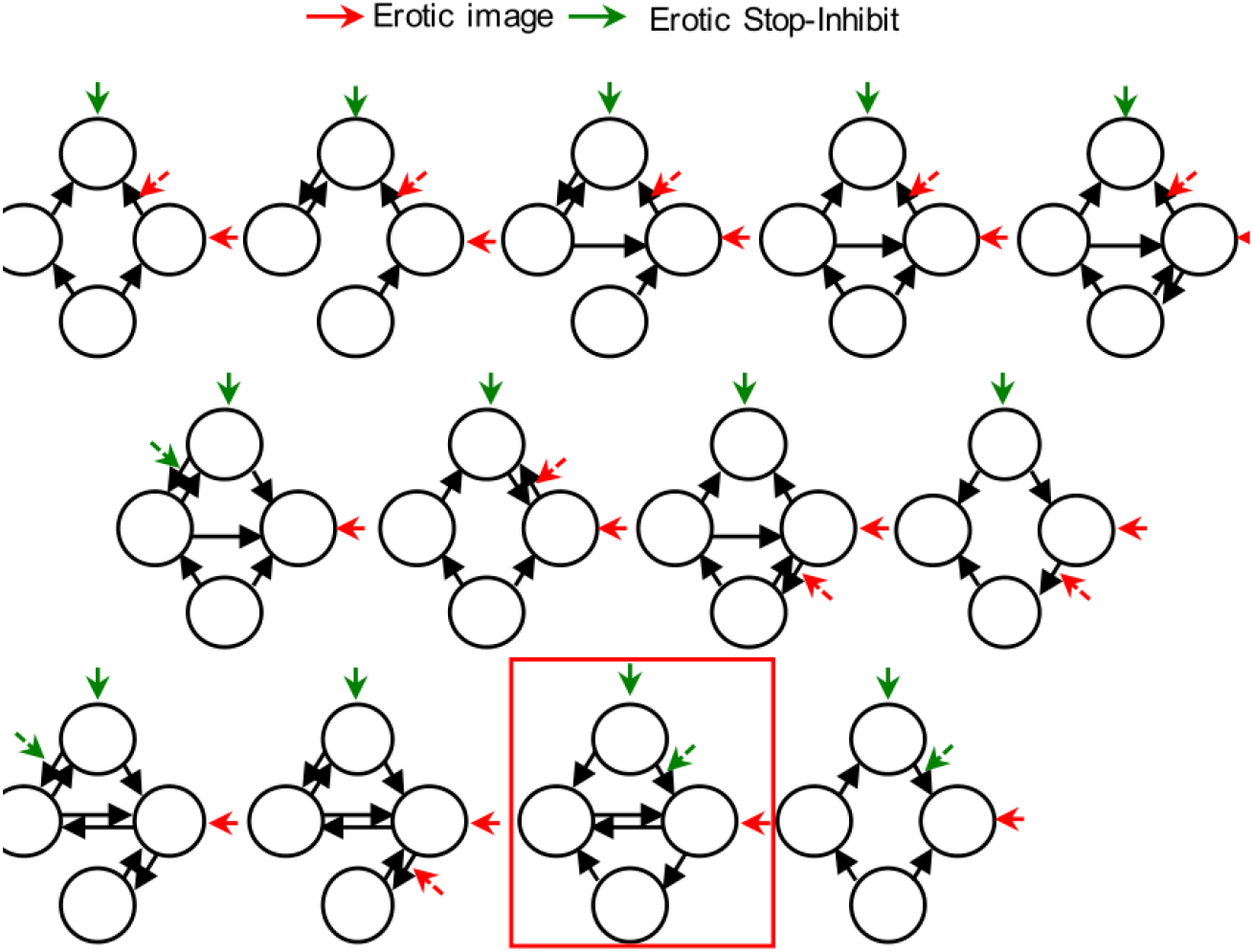
Structure of the 13 DCMs tested and selection of most likely model. The 13 generative models represent alternative hypotheses of cortico-subcortical interactions during cognitive control while exposed to sexual cues in hypersexuality. The most likely model after BMS is highlighted in red square.

**Figure S2.**
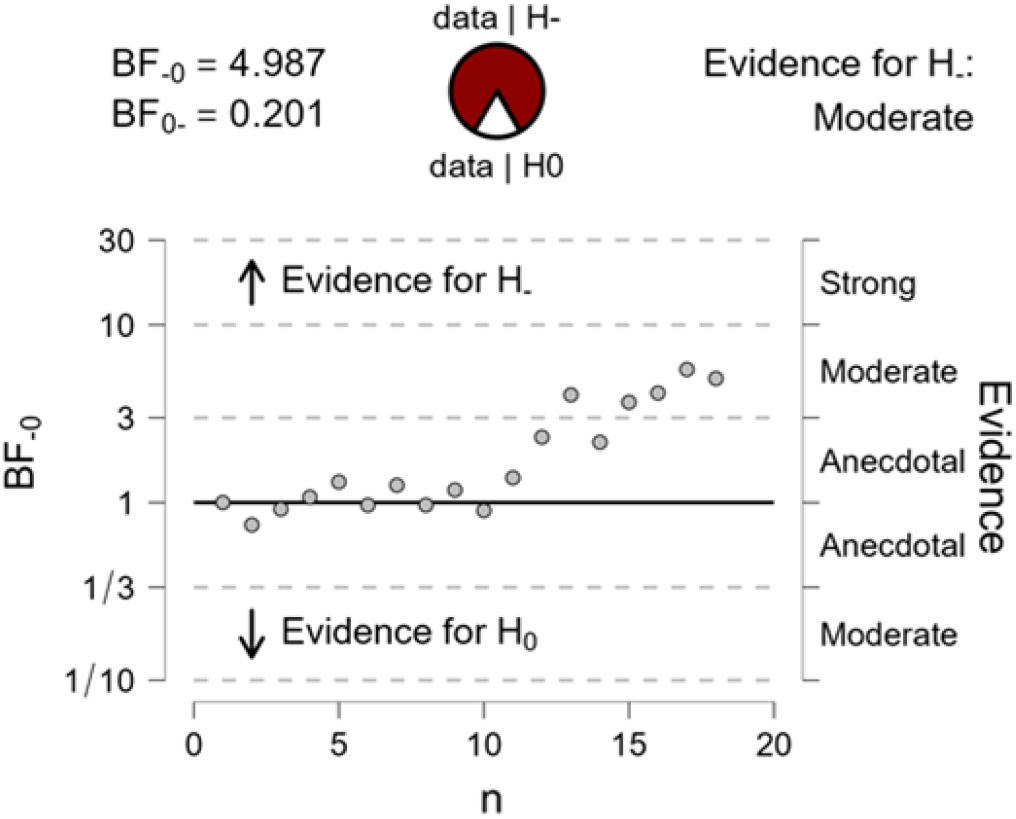
Bayesian factor sequential analysis for SSRT. Study 1 (PD+HS on < off).

**Figure S3.**
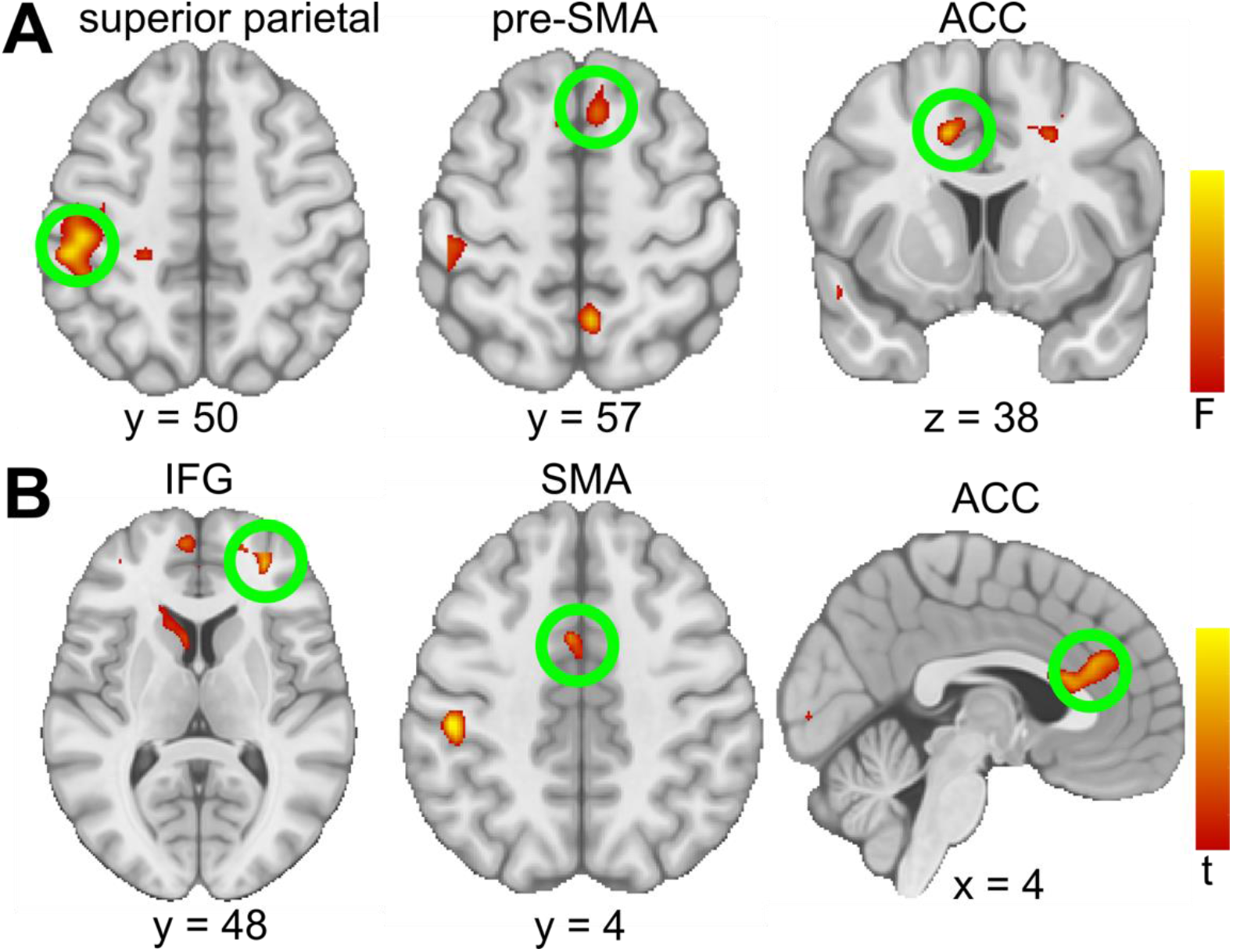
StopInhibit vs. StopFail activity maps. **(A)** Main effect of medication and **(B)** activity driven by PD+HS patients (on vs off medication). IFG = Inferior-frontal gyrus.

**Figure S4.**
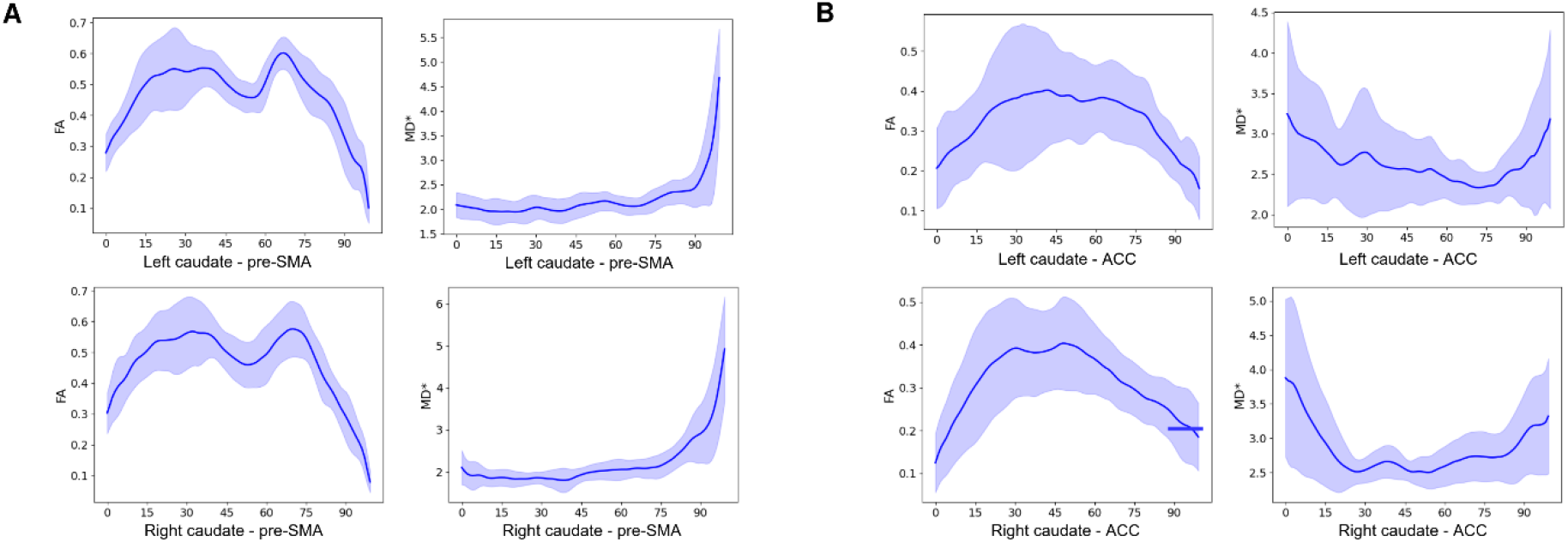
Tracts representation and DTI scalar measures (FA, MD*) for controls. **(A)** Fractional anisotropy (FA) and mean diffusivity (MD) along the segments of right-left caudate and pre-SMA tracts and **(B)**, along the segments of right-left caudate and anterior cingulate cortex tracts.

**Figure S5.**
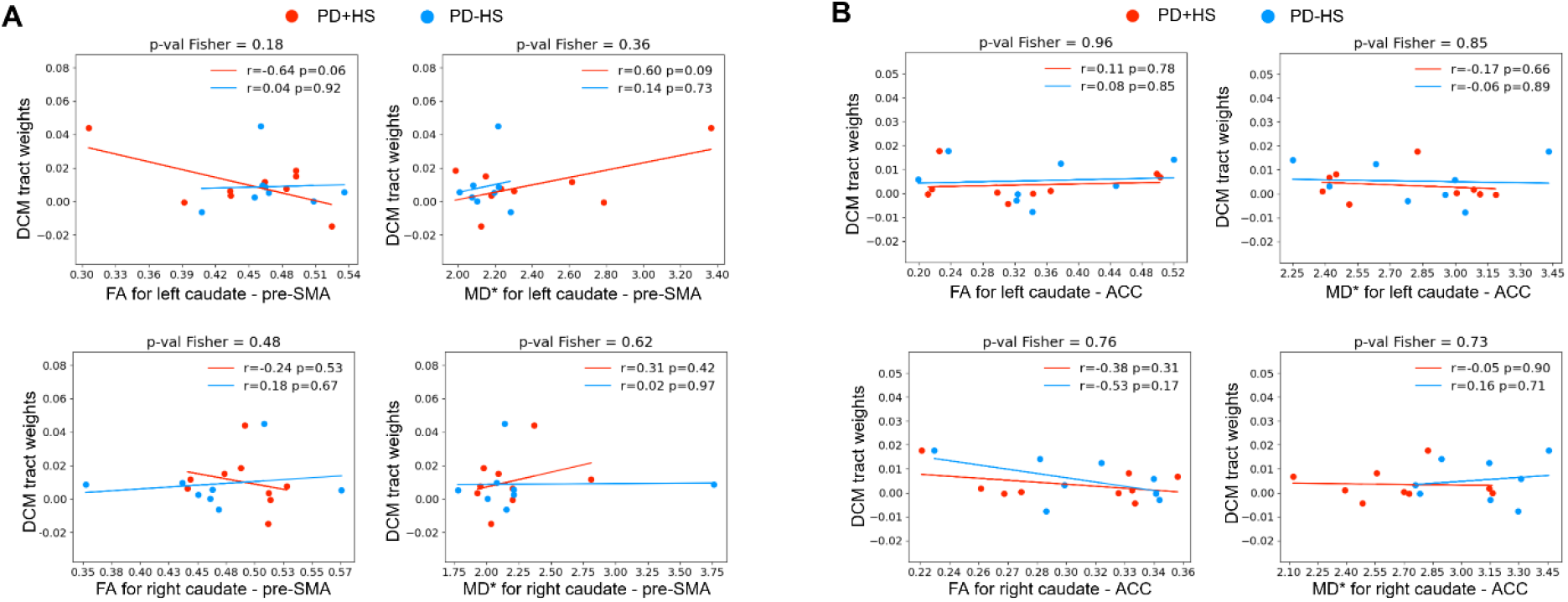
Correlation between diffusion tensor imaging (DTI) scalar measures – fractional anisotropy (*FA*), mean diffusivity (*MD*\*) – and *SSRT*. Comparisons included PD+HS (n=10) and PD-HS patients (n=14). **(A)** Correlation between FA and SSRT; **(B)** Correlation between MD* and SSRT (MD* is also called trace or TR, which is MD*3).

**Figure S6.**
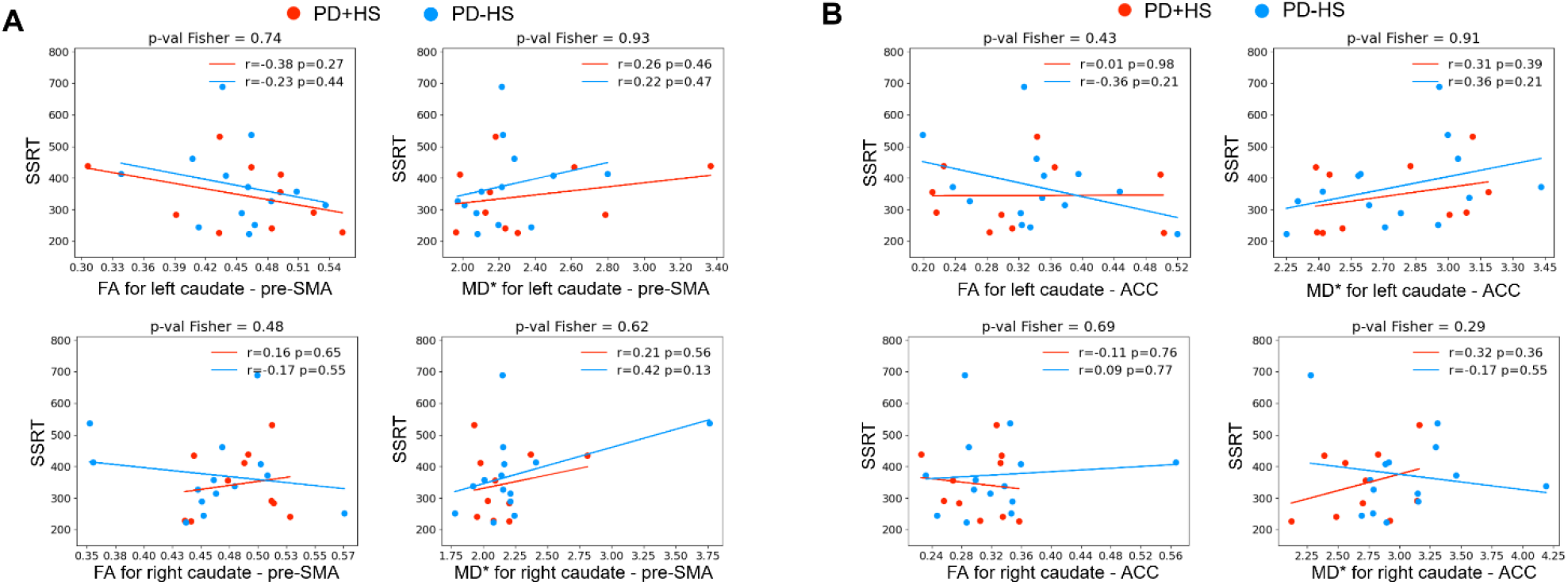
Correlation between DTI scalar measures (FA, MD*) and DCM parameters. Comparisons included PD+HS (n=9) and PD-HS patients (n=8). (A) Correlation between FA and DCM parameters; (B) Correlation between MD* and DCM parameters (MD* is also called trace or TR, which is MD*3).

**Figure S7.**
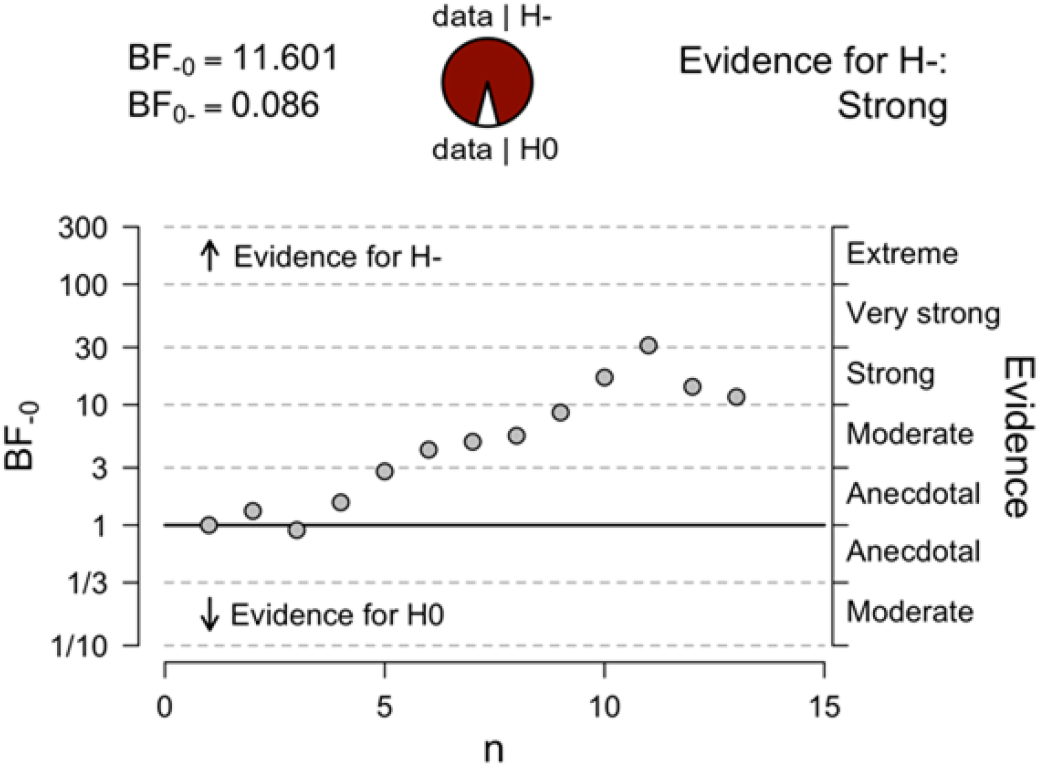
Bayesian factor sequential analysis for SSRT. Study 2 (real < sham).

**Table S1.**
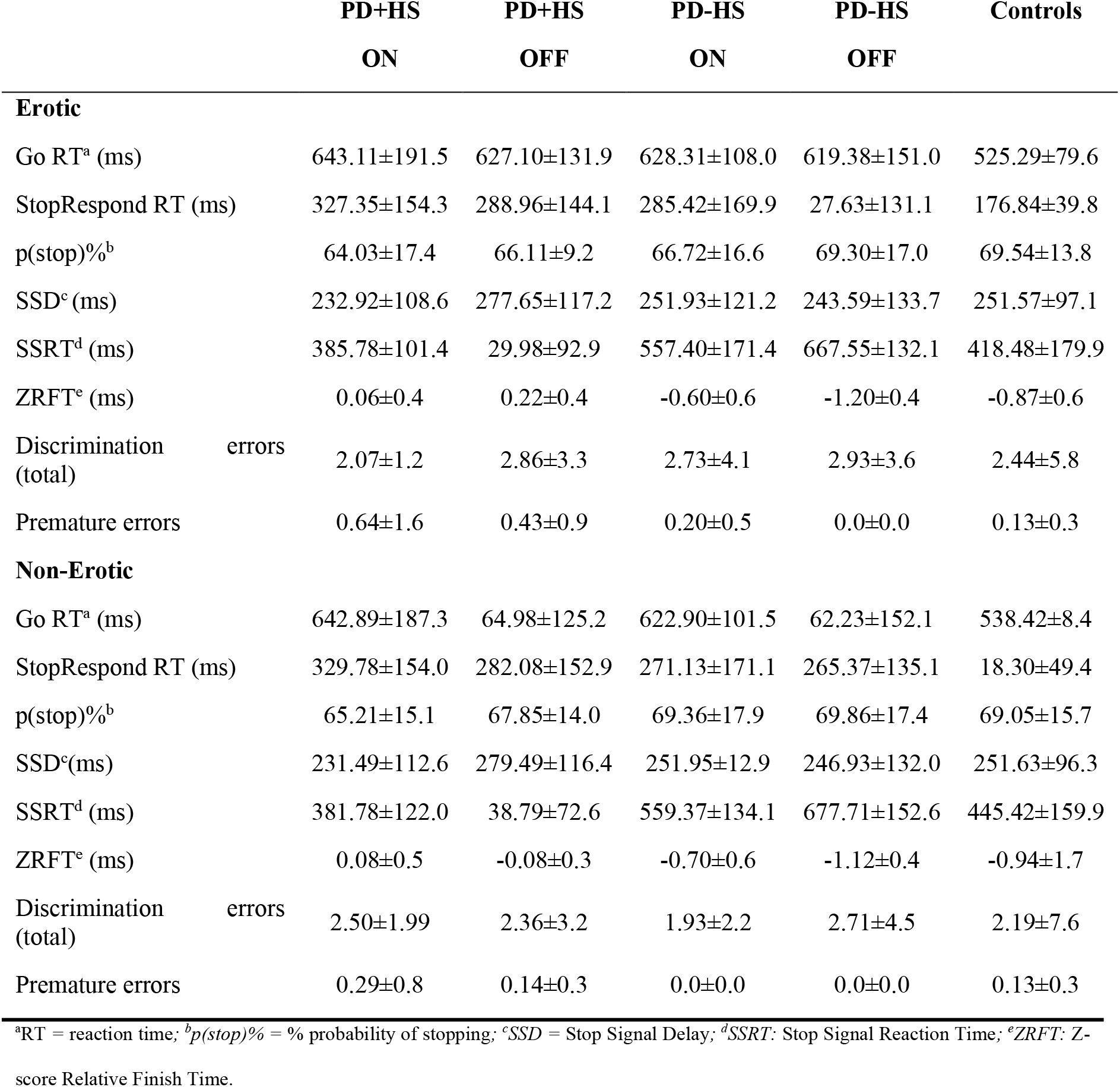
Erotic stop signal reaction time results per *Medication* and *Group* conditions.

**Table S2.** Stop-Inhibit vs. Go trials in the erotic condition.

| Tests and anatomical structures | X | Y | Z | T-value |
| --- | --- | --- | --- | --- |
| **Group** |  |  |  |  |
| *Motor cortex* | 20 | -36 | 60 | 5.71** |
| *Cerebellum* | 24 | -16 | 52 | 5.18** |
|  | 20 | -44 | -28 | 4.52** |
|  | 16 | -34 | -32 | 3.66** |
| **PD-HS > PD+HS off** |  |  |  |  |
| *Cingulate* | 18 | -10 | 46 | 5.77 |
| *Superior parietal lobe* | 26 | -24 | 50 | 4.91 |
| *Motor cortex* | 20 | -36 | 62 | 4.5 |

Tests and anatomical structures for the successful (*stop-inhibit*) vs *go* trials contrast in the erotic condition. $p < 0.05$ FWE cluster-wise corrected; \* $p < .005$ ; \*\* $p < .001$ .

**Table S3.** Stop Success vs Stop Fail under erotic condition.

| Tests and anatomical structures | X | Y | Z | T-value |
| --- | --- | --- | --- | --- |
| **Medication** |  |  |  |  |
| *Superior parietal lobe* | -44 | -24 | 46 | 4.18** |
|  | -50 | -30 | 50 | 3.86** |
|  | -46 | -38 | 52 | 3.41** |
| *Anterior cingulate cortex (ACC)* | -12 | 10 | 40 | 3.75* |
| *Pre-supplementary motor area (pre-SMA)* | 6 | 20 | 56 | 3.41* |
| *Supplementary motor area* | 0 | -8 | 70 | 3.20* |
| **PD+HS on > off** |  |  |  |  |
| *Frontal pole* | 16 | 62 | 18 | 4.44** |
| *Anterior cingulate cortex (ACC)* | 0 | 24 | 14 | 4.13** |
| *Inferior-frontal gyrus* | 30 | 46 | 10 | 4.06** |
| *Supplementary motor area* | -4 | 6 | 44 | 3.69* |
|  | -6 | 6 | 56 | 3.34* |
|  | 10 | 6 | 50 | 3.27* |
| **PD+HS > PD-HS on** |  |  |  |  |
| *Superior parietal lobe* | -44 | -24 | 46 | 4.73** |
|  | -50 | -30 | 50 | 4.34** |
|  | -46 | -38 | 52 | 3.81** |
| *Pre-supplementary motor area (pre-SMA)* | -12 | 10 | 40 | 4.21** |
|  | 6 | 20 | 56 | 3.81** |
|  | -24 | 14 | 62 | 3.59* |

Tests and anatomical structures for the successful (*stop-inhibit*) vs unsuccessful (*failed*) stop trials contrast in the erotic condition. $p < .05$
FWE cluster-wise corrected; \* $p < .005$ ; \*\* $p < .001$ .

**Table S4.** Stop fail in Erotic vs. Non-erotic trials.

| Tests & Anatomical structures | X | Y | Z | T-value |
| --- | --- | --- | --- | --- |
| **Medication x Group** |  |  |  |  |
| *Superior parietal lobe* | -44 | -28 | 46 | 4.27** |
| *Premotor area (BA6)* | -36 | -12 | 64 | 3.85* |
| *Supplementary motor area (Pre-SMA; BA6)* | 0 | -6 | 68 | 3.31* |
| *Superior frontal gyrus (BA6)* | -16 | 2 | 68 | 3.26* |
| **PD+HS on > off** |  |  |  |  |
| *Caudate* | -4 | 8 | 8 | 4.16*^ |
|  | -4 | 6 | 18 | 3.51*^ |
|  | -12 | 2 | 14 | 3.18*^ |
| **PD+HS > PD-HS on** |  |  |  |  |
| *Superior parietal lobe (BA31)* | -52 | -20 | 52 | 4.11** |
| *Caudate* | -10 | -4 | 20 | 3.92* |
|  | -10 | 8 | 14 | 3.65* |
|  | -4 | 10 | 6 | 3.56* |

Tests and anatomical structures for unsuccessful stop (*stop-fail*) in erotic vs. non-erotic trials, $p < .05$ FWE cluster-wise corrected;
\* $p < .005$ ; \*\* $p < .001$ .

**Table S5.** Behavioral results of the stop signal task (study 2) after real and sham intermittent theta burst stimulation.

|  | Real | Sham | *p*-value | Real | Sham | *p*-value |
| --- | --- | --- | --- | --- | --- | --- |
|  | Erotic | Erotic |  | Non-erotic | Non-erotic |  |
| Go RT<sup>a</sup> (ms) | 564.82±106.38 | 593.96±115.52 | .06 | 552.00±93.11 | 581.68±105.52 | .05 |
| StopRespond<br>RT<sup>a</sup> (ms) | 213.05±90.92 | 242.50±106.40 | .11 | 222.71±109.09 | 221.95±106.88 | .96 |
| p(stop)%<sup>b</sup> | 71.37±14.56 | 70.50±19.11 | .72 | 71.17±13.78 | 71.20±17.70 | .99 |
| SSD<sup>c</sup> (ms) | 304.99±128.79 | 303.31±137.56 | .89 | 297.72±118.86 | 307.27±136.37 | .47 |
| SSRT (ms) | 341.51±138.42 | 392.47±155.06 | .02 | 352.12±182.19 | 398.13±204.05 | .04 |
| ZRFT (ms) | -0.56±0.54 | -0.56±0.63 | .98 | -0.58±0.66 | -0.63±0.69 | .59 |
| Discrimination<br>errors (total) | 2.89±3.53 | 2.50±3.24 | .62 | 3.33±4.46 | 2.83±3.93 | .61 |

Comparisons of task measures between real and sham sessions for erotic and non-erotic trials (*p* value: paired t-tests). <sup>a</sup>RT = reaction time;
<sup>b</sup>p(stop)% = % probability of stopping; <sup>c</sup>SSD = Stop Signal Delay; <sup>d</sup>SSRT: Stop Signal Reaction Time; <sup>e</sup>ZRFT: Z-score Relative Finish Time.

